# Investigating the relationship between extreme weather and cryptosporidiosis and giardiasis in Colorado: a multi-decade study using distributed-lag nonlinear models

**DOI:** 10.1101/2023.08.31.23294911

**Authors:** Elise N. Grover, James L. Crooks, Elizabeth J. Carlton, Sara H. Paull, William B. Allshouse, Rachel H. Jervis, Katherine A. James

## Abstract

Environmentally-mediated protozoan diseases like cryptosporidiosis and giardiasis are likely to be highly impacted by extreme weather, as climate-related conditions like temperature and precipitation have been linked to their survival, distribution, and overall transmission success. Our aim was to investigate the relationship between extreme temperature and precipitation and cryptosporidiosis and giardiasis infection using monthly weather data and case reports from Colorado counties over a twenty-one year period. Data on reportable diseases and weather among Colorado counties were collected using the Colorado Electronic Disease Reporting System (CEDRS) and the Daily Surface Weather and Climatological Summaries (Daymet) Version 3 dataset, respectively. We used a conditional Poisson distributed-lag nonlinear modeling approach to estimate the lagged association (between 0 and 12-months) between relative temperature and precipitation extremes and the risk of cryptosporidiosis and giardiasis infection in Colorado counties between 1997 – 2017, relative to the risk found at average values of temperature and precipitation for a given county and month. We found distinctly different patterns in the associations between temperature extremes and cryptosporidiosis, versus temperature extremes and giardiasis. When maximum or minimum temperatures were high (90^th^ percentile) or very high (95^th^ percentile), we found a significant increase in cryptosporidiosis risk, but a significant decrease in giardiasis risk, relative to risk at the county and calendar-month mean. Conversely, we found very similar relationships between precipitation extremes and both cryptosporidiosis and giardiasis, which highlighted the prominent role of long-term (>8 months) lags. Our study presents novel insights on the influence that extreme temperature and precipitation can have on parasitic disease transmission in real-world settings. Additionally, we present preliminary evidence that the standard lag periods that are typically used in epidemiological studies to assess the impacts of extreme weather on cryptosporidiosis and giardiasis may not be capturing the entire relevant period.

## 1. Introduction

In recent decades, rapid increases in global temperatures are giving rise to an array of changes in local ecosystems, local climates and the frequency and magnitude of extreme weather, all of which are likely to impact environmentally-mediated illnesses such as those transmitted by the parasitic protozoa, *Cryptosporidium* and *Giardia* (Fletcher, Stark et al. 2012, Luber and Lemery 2015, Pozio 2020, IPCC 2021). These protozoan diseases are a leading cause of the estimated 1.7 billion cases of diarrheal disease that occur every year (Lane and Lloyd 2002, Fayer, Dubey et al. 2004, Chalmers and Davies 2010, Liu, Platts-Mills et al. 2016, Efstratiou, Ongerth et al. 2017), with an estimated 33,900 deaths and 2.94 million disability-adjusted life-years (DALYs) lost to illness caused by enteric protozoa annually (Torgerson, Devleesschauwer et al. 2015). *Cryptosporidium* and *Giardia* are both zoonotic, waterborne parasites that are transmitted by animals and humans via the fecal-oral route, and are most frequently acquired by humans from contaminated drinking or recreational water, though contact with livestock and livestock-contaminated agricultural products can also play a significant role in human transmission (Hunter and Thompson 2005, Karanis, Kourenti et al. 2006, Budu-Amoako, Greenwood et al. 2011, Dumètre, Aubert et al. 2012, Dreelin, Ives et al. 2014, Painter, Gargano et al. 2016, Ligda, Claerebout et al. 2020). As such, regions of the world that have limited access to clean water, are prone to water scarcity, or lack infrastructure to support widespread access to improved sanitation tend to be the most at risk of cryptosporidiosis and giardiasis (Hunter and Thompson 2005, Ahmed, Guerrero Flórez et al. 2018). Nevertheless, high income settings can experience increases in protozoan enteric disease risk as a result of societal factors like rising irrigation dependence (Liu, Folberth et al. 2013, Lake and Barker 2018), growing participation in outdoor recreational activities (Hlavsa, Roberts et al. 2015, Painter, Gargano et al. 2016, Association 2021, Ferguson, McIntosh et al. 2022), and environmental phenomena such as the rising frequency and severity of extreme temperatures and precipitation (Fletcher, Stark et al. 2012, Molloy, Dreelin et al. 2017, Intergovernmental Panel on Climate Change 2018, Akhtar 2020, IPCC 2021). Given the high burden of environmentally-mediated illnesses like cryptosporidiosis and giardiasis, as well as uncertainty that arises around rapidly changing climate systems, investigations into the relationship between local transmission dynamics and climate extremes are of paramount importance.

While several studies have demonstrated associations between climate conditions and cryptosporidiosis and giardiasis, these relationships can vary drastically by local context (e.g., local climate and ecosystem type, season, land use, water regulations, etc.,) (Naumova, Jagai et al. 2007, Fletcher, Stark et al. 2012, Lal, Hales et al. 2012, Semenza, Herbst et al. 2012). For instance, a meta-analysis conducted by Jagai and colleagues (2009) demonstrated that while temperature was more strongly associated with cryptosporidiosis in temperate climates, precipitation was a stronger predictor in tropical climates, though notably, neither temperature nor precipitation was significantly associated with cryptosporidiosis in arid or semi-arid climates (Jagai, Castronovo et al. 2009). In the case of temperature and cryptosporidiosis, more studies have reported a positive relationship between cryptosporidiosis and temperature (Hu, Tong et al. 2007, Hu, Tong et al. 2007, Naumova, Jagai et al. 2007, Lake, Pearce et al. 2008, Jagai, Castronovo et al. 2009, Ajjampur, Liakath et al. 2010, Hu, Mengersen et al. 2010, Hu, Mengersen et al. 2010, Kent, McPherson et al. 2015, Ikiroma and Pollock 2021, Ma, Destouni et al. 2021, Wang, Wang et al. 2023) than a negative one (Britton, Hales et al. 2010, Kent, McPherson et al. 2015). In the case of giardiasis, the reported relationship with temperature has been less consistent (Wang, Wang et al. 2023), with some studies suggesting a positive association (Naumova, Jagai et al. 2007, Britton, Hales et al. 2010), others reporting a negative one (Wilkes, Edge et al. 2011, Li, Chase et al. 2019, Masina, Shirley et al. 2019), and at least one suggesting no association (Lal, Ikeda et al. 2013). Likewise, studies assessing the relationship between precipitation and cryptosporidiosis and giardiasis have had similarly mixed results (Miller, Lewis et al. 2007, Naumova, Jagai et al. 2007, Keeley and Faulkner 2008, Mons, Dumètre et al. 2009, Britton, Hales et al. 2010, Gonzalez-Moreno, Hernandez-Aguilar et al. 2013, Lal, Baker et al. 2013, Daniels, Smith et al. 2016, Liu, Gong et al. 2020, Wang, Wang et al. 2023).

Further challenging our understanding of how rapidly changing climates can impact cryptosporidiosis and giardiasis transmission is the fact that to date, few studies have specifically assessed local climate extremes in relation to cryptosporidiosis and giardiasis (Carlton, Woster et al. 2016). This is particularly true in the case of temperature extremes, where the vast majority of the literature comes from experimental, rather than epidemiological studies. Overall, experimental studies have highlighted that both *Cryptosporidium* oocysts and *Giardia* cysts can survive and remain viable after short-term temperature extremes (Wickramanayake, Rubin et al. 1985, Fayer and Nerad 1996, Fayer, Trout et al. 1998, Fayer, Morgan et al. 2000, King, Keegan et al. 2005, Alum, Absar et al. 2014). For example, *Cryptosporidium* oocysts stored in water held at -10°C and -5°C remained infectious for up to 1 and 8 weeks of storage respectively, while those held at 35°C for up to a week of storage could produce light infections in mice (Fayer and Nerad 1996, Fayer, Trout et al. 1998). Meanwhile, *Giardia* cysts are more sensitive to temperatures below freezing or above 30°C than *Cryptosporidium* oocysts, with one study finding low viability (<0.2%) when *Giardia* cysts were stored at either -6°C or 37°C for more than 3 days (Wickramanayake, Rubin et al. 1985).

By contrast, few epidemiological studies have focused specifically on the relationship between temperature extremes and protozoan disease transmission in real-world settings. In a recent review by Wang, Wang & Cao (2023), the authors identified a total of three studies that assessed the relationship between extreme high temperatures – expressed as daily, weekly or monthly maximum temperature – and cryptosporidiosis, all of which used a lag of ≤3 months, and all found a positive association with maximum temperature in at least one study location (Hu, Mengersen et al. 2010, Kent, McPherson et al. 2015, Brankston, Boughen et al. 2018, Wang, Wang et al. 2023). Meanwhile, the same review (Wang, Wang et al. 2023) identified just one study that assessed the relationship between low temperature – specifically, mean minimum monthly temperature – and cryptosporidiosis, which highlighted positive association between 0-, 1-, and 2-month lagged mean minimum monthly temperature and cryptosporidiosis in metropolitan areas, but a negative relationship between 3-month lagged minimum monthly temperature and cryptosporidiosis in rural areas (Kent, McPherson et al. 2015).

Precipitation extremes have been more frequently studied in relation to cryptosporidiosis and giardiasis transmission (Keeley and Faulkner 2008, Chhetri, Takaro et al. 2017, Lal and Konings 2018, Brunn, Fisman et al. 2019, Chhetri, Galanis et al. 2019, Forbes, Hosking et al. 2021, Ma, Destouni et al. 2021, Graydon, Mezzacapo et al. 2022), though the magnitude of the relationships tend to be highly dependent on background conditions, and subsequently, how variables and lags are defined and measured. Both periods of drought and heavy rainfall can cause inefficiencies and reduced effectiveness of water treatment systems, which can subsequently impact the risk of cryptosporidiosis and giardiasis transmission (Curriero, Patz et al. 2001, Thomas, Charron et al. 2006, Nichols, Lane et al. 2009, Luber and Lemery 2015, Chhetri, Takaro et al. 2017, Schreiber, Heinkel et al. 2019). For example, studies conducted in Australia and the Netherlands have found that periods of drought were associated with decreased water quality (Senhorst and Zwolsman 2005, Zwolsman and van Bokhoven 2007), and an increased risk of cryptosporidiosis (Lal and Konings 2018). Likewise, periods of heavy rainfall can also cause sewage system overflow, contamination of irrigation systems, wells and private water supplies, and subsequent food and waterborne illnesses (Semenza, Herbst et al. 2012, Luber and Lemery 2015). For instance, researchers in Vancouver, Canada found that the risk of runoff-related cryptosporidiosis and giardiasis incidence was highest when a period of excessive dryness was followed by heavy rainfall (Chhetri, Takaro et al. 2017). Exposure via runoff and waste water system failures may be particularly important for parasitic protozoan diseases, which are more resilient than most bacteria and viruses in the face of conventional water treatment methods (Omarova, Tussupova et al. 2018), and are less commonly the target of operational surveillance systems (Fletcher, Stark et al. 2012).

The majority of studies assessing the relationship between temperature and precipitation extremes and cryptosporidiosis and giardiasis have assessed lag periods of between climate and disease of up to three months (Hu, Tong et al. 2007, Naumova, Jagai et al. 2007, Hu, Mengersen et al. 2010, Hu, Mengersen et al. 2010, Kent, McPherson et al. 2015, Chhetri, Takaro et al. 2017, Brankston, Boughen et al. 2018, Brunn, Fisman et al. 2019, Chhetri, Galanis et al. 2019), however, we know that pathogen survival in the ambient environment is likely longer under optimal conditions. While determining the maximum survival time in the ambient environment can be challenging, controlled studies have demonstrated that *Giardia* cysts submerged in river and lake water at winter temperatures maintained viability at 12-weeks post-baseline (deRegnier, Cole et al. 1989), while *Cryptosporidium* oocyst infectivity can remain after being stored in water for over five months at 5-15°C (Fayer, Trout et al. 1998), and up to a year when stored in low turbidity water (Badenoch 1990). Given their demonstrated capacity for prolonged survival, epidemiological studies that assess the potential for longer lagged effects in real-word settings are needed.

Table 1 provides examples from the literature and a summary of some of the hypothesized mechanisms by which extreme precipitation and temperature may impact *Cryptosporidium* and *Giardia* mobilization and survival and subsequent cryptosporidiosis and giardiasis transmission. In light of the many influences that weather and the environment have on the transmission of protozoan pathogens, as well as remaining uncertainty surrounding the time- and location-specific conditions that may amplify or reduce risk, localized investigations of weather extremes and their lagged effects on transmission are needed.

**Table 1.** Hypothesized mechanisms by which temperature and precipitation extremes can impact protozoan disease transmission.

| **Weather extremes** | **Direction of association** | **Examples of hypothesized impacts on pathogens survival, mobilization and distribution** | **Examples of hypothesized climate-related pathogen exposure routes** |
| --- | --- | --- | --- |
| **Temperature** |  |  |  |
| Above average temperature | Increase in protozoan transmission | • Prolonged infectivity of oocysts (King and Monis 2007)<br>• Enhanced survivability of temperature-dependent mechanical vectors (Conn, Weaver et al. 2007) | • Changes in outdoor recreational activities and water quality (Schets, Husman et al. 2011, Young, Sanchez et al. 2022)<br>• Changes in livestock roaming, health, and potential for direct or indirect livestock contact (Thornton, van de Steeg et al. 2009)<br>• Changes in drinking water supply and quality (Boithias, Choisy et al. 2016) |
|  | Decrease in protozoan transmission | • Pathogen desiccation and decreased viability (Dixon 2014)<br>• Accelerated degradation of oocysts and cysts (Olson, Goh et al. 1999) |  |
| Below average temperature | Increase in protozoan transmission | • Prolonged survival on surfaces (Alum, Absar et al. 2014)<br>• Prolonged survival in water (deRegnier, Cole et al. 1989)<br>• Prolonged infectivity in warm seasons (Fayer, Trout et al. 1998) |  |
|  | Decrease in protozoan transmission | • Decreased survival in cold seasons (Fayer, Trout et al. 1998) |  |
| **Precipitation** |  |  |  |
| Above average precipitation | Increase in protozoan transmission | • Increase in contaminated runoff (Davies-Colley, Nagels et al. 2008)<br>• Sewage overflows (Tolouei, Dewey et al. 2019) | • Changes in outdoor recreational water activities and water quality (Gunady, Koutsoukos et al. 2016, Iniguez-Armijos, Sánchez et al. 2020)<br>• Changes in farmland irrigation practices and runoff (Chaidez, Soto et al. 2005, Spanakos, Biba et al. 2015, Amanullah, Khalid et al. 2020)<br>• Changes in drinking water supply and quality (Boithias, Choisy et al. 2016)<br>• Changes in potential for direct contact with contaminated water (Tolouei, Dewey et al. 2019) |
|  | Decrease in protozoan transmission | • Dilution of overall pathogen concentration (Ajeagah, Njine et al. 2010) |  |
| Below average precipitation | Increase in protozoan transmission | • Increase in effluent concentrations in surface or stored water supplies (Boxall, Hardy et al. 2009, Boithias, Choisy et al. 2016) |  |
|  | Decrease in protozoan transmission | • Decreased water turbidity and pathogen contamination (Chhetri, Takaro et al. 2017, De Roos, Gurian et al. 2017)<br>• Decrease in contaminated runoff (Miller, Lewis et al. 2008) |  |

In this study, we investigated the potential impact of temperature and precipitation extremes on cryptosporidiosis and giardiasis risk in Colorado counties between 1997 – 2017. Our approach was designed to allow evaluation of lagged relationships up to 12 months, addressing a gap in the epidemiological evidence to date. We used a flexible, distributed-lag nonlinear modeling approach to explore nonlinear outcome-predictor relationships and permit lagged effects to be spread over time, two characteristics that are desirable when exploring the impacts of highly variable environmental conditions on health outcomes. In so doing, we provide valuable insight on the relationships between real-world shifts in temperature and precipitation and two important, but often overlooked sources of waterborne illness, with the goal of advancing our basic understanding of the mechanisms by which climate impacts infectious diseases and improving risk forecasting.

## 2. Materials and Methods

### 2.1. Study design

In this study, we assessed the impacts of precipitation and temperature extremes on reported cases of cryptosporidiosis and giardiasis in Colorado counties between 1997 and 2017. Specifically, our primary objective was examining the effects of extreme temperature and precipitation values – defined as the 5^th^, 10^th^, 90^th^ and 95^th^ percentile values of countywide monthly averages for maximum daily temperature (TMAX), minimum daily temperature (TMIN) and total precipitation (PRECIP) (each mean-centered by county and calendar month) – on monthly counts of reported human cryptosporidiosis and giardiasis cases. We also investigated the relevant lag periods over which extreme weather was associated with cryptosporidiosis and giardiasis in Colorado counties and evaluated whether season served as an effect modifier within our models. We used a distributed lag nonlinear modeling (DLNM) approach, conditioned on total case counts within each of the twelve calendar-months for a given county (Gasparrini, Armstrong et al. 2010, Armstrong, Gasparrini et al. 2014). The Colorado Multiple Institution Review Board reviewed and approved of this study as meeting Institutional Review Board exemption status.

### 2.2. Cryptosporidiosis and giardiasis incidence

Data on reportable diseases in Colorado were collected using the Colorado Electronic Disease Reporting System (CEDRS), which is made available upon request from the Colorado Department of Public Health and Environment (CDPHE). Our study spanned the 21-year period between January 1997 through December 2017, using monthly case reports of cryptosporidiosis and giardiasis by county as our two outcomes of interest. As both cryptosporidiosis and giardiasis are mandatory reportable diseases and CDPHE conducted lab audits to ensure proper monthly reporting, for the purposes of this study we assumed that cases of sufficient severity to warrant medical attention and appropriate testing in Colorado counties between 1997 and 2017 were captured in the CEDRS dataset.

In order to estimate incidence, we used annual county population estimates from the Colorado Department of Local Affairs (Colorado Department of Local Affairs 2019). Although Broomfield County was approved by voters as Colorado’s 64^th^ county in 1998, it was not included as its own county in this analysis until 2000, as prior to this point, the Colorado Department of Local Affairs census data recorded Broomfield’s population as a part of Boulder County’s population.

### 2.3. Climatological variables

For our climatological variables, we used the Daily Surface Weather and Climatological Summaries (Daymet) Version 3 dataset derived by the National Center for Atmospheric Research (NCAR), which provides daily estimates of weather parameters across a 1-kilometer grid spacing for North America (Thornton, Thornton et al. 2017). In order to align the resolution of our climatological data with the monthly resolution of case report data that was available for all Colorado counties between 1997 – 2017, TMAX (°C) and TMIN (°C) were each averaged across all 1-km grid points within a given county for each month between January 1996 and December 2017, such that each county-month had a single estimated average value for TMIN and TMAX. PRECIP was estimated for each county-month using total daily precipitation (mm) measured at each 1-km grid point within a county’s borders, summed across all days in each month and averaged across all grid points in the county. The Daymet Version 3 dataset includes all forms of precipitation (e.g., snow, sleet, rain), which are converted to a water-equivalent depth in millimeters. Notably, while our outcome data begins in 1997, the data for the explanatory variables begins in January 1996 to allow for up to 12-month lagged effects for each of our weather parameters.

To make the definition of extremes most relevant to the local weather conditions found in Colorado counties, we mean-centered our weather variables by county and calendar-month, such that values could be interpreted as hot/cold or wet/dry for the specific location and time of year. All three weather variables were subsequently mean-centered by county and calendar-month, allowing TMAX, TMIN and PRECIP to be defined relative to their location and time. For example, the average TMAX was 8.9°C for Adams County in December 2017, while the mean-centered (MC)-TMAX was +2.6°C (as compared to other Decembers between 1996 – 2017 in Adams County), indicating a warmer than average December in Adams County in 2017.

In this analysis, we were ultimately interested in the association between extreme temperature and precipitation and cryptosporidiosis and giardiasis incidence. Thus, to define the set of relevant predictor values to be evaluated as the numerator in our relative risk calculations, we used the 5^th^, 10^th^, 90^th^ and 95^th^ percentile values of our mean-centered weather variables, referred to respectively as very low, low, high and very high throughout this analysis, as detailed in Appendix Table A1.

### 2.4. Analysis

To assess the lagged effects of MC-TMAX, MC-TMIN and MC-PRECIP on human cryptosporidiosis and giardiasis cases in Colorado counties between 1997-2017, we used a conditional Poisson DLNM approach (Gasparrini, Armstrong et al. 2010, Armstrong, Gasparrini et al. 2014). We investigated a wide range of potential lags between 0- and 12-months by varying the number of internal knots (0-6) for the spline functions used to define the lag space and comparing model performance across these formulations of the lag space. Whereas 2- and 4-month lags were selected because they aligned with the typical lag range indicated in the literature (Hu, Tong et al. 2007, Naumova, Jagai et al. 2007, Hu, Mengersen et al. 2010, Hu, Mengersen et al. 2010, Kent, McPherson et al. 2015, Ikiroma and Pollock 2021), 6-, 8-, 10- and 12-month lags was also examined in detail in order to determine whether extreme weather could have more prolonged impacts of on giardiasis or cryptosporidiosis case counts in Colorado counties between 1997 – 2017. Ultimately, this design allowed us to identify potential longer-term trends that might have otherwise been missed if only short-term lags were assessed. The impacts of MC-TMAX, MC-TMIN and MC-PRECIP on monthly cryptosporidiosis and giardiasis county case counts were each assessed independently, making a total of six outcome-predictor combinations.

The steps that were taken to conduct this analysis for each of the six outcome-predictor pairs of interest to this study are outlined in Figure 1. In Step 1, initial exploratory analyses were conducted to summarize between-season differences in our outcome-predictor pairs across our study period. A three-month definition of season was also used in our exploratory assessment to investigate the distribution of the data across the calendar year. Winter was defined as December – February; spring as March – May; summer as June – August; and fall as September – November.

**Figure 1.**
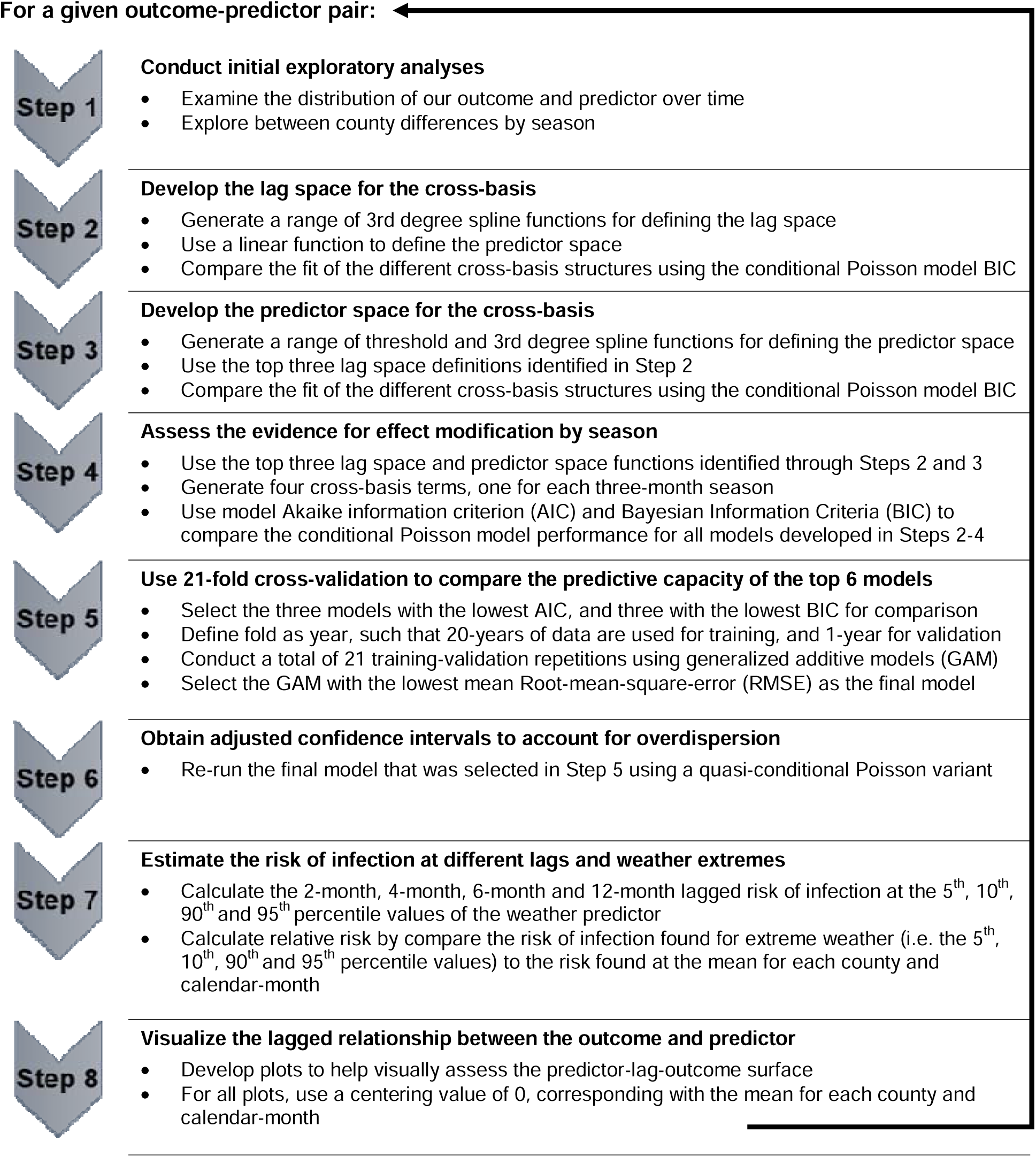
Illustration of the analytical steps used in this study. **Figure 1 caption:** The 8-Step analytical process that was used for each outcome and predictor of interest in this study is outlined above. The two outcomes in this study were case counts of cryptosporidiosis, and case counts of giardiasis. The three predictors were the average maximum monthly temperature, the average minimum monthly temperature, and the total monthly precipitation. We therefore repeated Steps 1-8 a total of six times for this study, once for each outcome-predictor pair.

Next, we developed several different cross-basis structures to investigate potential non-linearities in the lagged predictor-outcome relationship over time and space, using the “dlnm” package in R (Gasparrini 2011, RStudio Team 2020). A cross-basis is a bi-dimensional space of functions that encapsulates both the shape of the relationship between the outcome and predictors (termed the “predictor space”), as well as the distributed lagged effects (i.e. the “lag space”) (Gasparrini, Armstrong et al. 2010). To develop a cross-basis with suitable functions defining both the predictor space and the lag space, we investigated the fit of different third-degree spline functions (natural spine, B-spline and penalized spline) with incrementally increasing degrees of freedom (maximum of seven), while the predictor space was held constant using a simple linear basis function. In all models, the maximum lag was defined as 12-months. The default settings for knot placements were used for each spline function such that internal knots were placed at equally spaced points along the lag-space, while the boundary knots were set to 0- and 12-months for the natural and B-spline functions. We used a conditional Poisson generalized nonlinear modeling (GNM) approach to compare model fit across the different lag space functions used in the cross-basis. All models also included an intercept, a continuous year term to account for linear time trends, an offset for county population (i.e., the natural log of the county’s annual population), and county-month as the conditioning (i.e., stratum) variable. Each model’s Bayesian information criterion (BIC) was used to identify the three best lag space definitions for the cross-basis.

Once suitable functions were identified for the lag space, Step 3 was to develop the predictor space using a similar process for the lag space. That is, the fit of different third-degree spline functions with incrementally increasing degrees of freedom (up to a maximum of five) were compared for the predictor space, while we defined the lag space using the top three previously identified lag space formulations. In addition to assessing different variations of third-degree spline functions, threshold functions were also assessed for the predictor space. The BIC value was used again to select the three best fit models across all the different cross-basis formulations.

We looked for evidence of potential effect modification by season in Step 4. Four separate cross-basis terms (one for each three-month season) were generated, using the top performing predictor space and lag space definitions previously identified through Steps 2 and 3. In total, the procedure outlined in steps 2-4 yielded a set of 50 models for each outcome-predictor pair, which were then compared a final time using both the Akaike information criterion (AIC) and BIC values to select a final subset of six models that were reserved for further exploration. To demonstrate the analytical process that was used to build suitable cross-basis structures in Steps 2-4, Appendix Table A2 provides an example from one outcome-predictor pair, detailing all model variants that were developed for MC-TMIN and cryptosporidiosis.

The predictive capacity of each of the six AIC/BIC-selected models for a given outcome-predictor was then assessed using cross-validation in Step 5. For each of the six models, repeated training and testing was conducted using a 21-fold cross-validation (Kuhn 2008). Each fold contained a year of observations, such that, within each fold, 20 years of data were used to train the model while the remaining year of county-month observations was used for validation. This training-validation process was repeated a total of twenty-one times, once for each year between 1997 and 2017. The mean RMSE value summarizing the performance across all tuning and validation iterations was then used as an indicator of the overall predictive skill of the model. The model with the lowest mean RMSE was selected as the final model (Appendix Table A3).

The final model was re-run using a conditional quasi-Poisson GNM variant to obtain adjusted confidence intervals to account for overdispersion (Step 6). As was done with for the conditional Poisson models, the quasi-conditional Poisson models included an intercept outside of the cross-basis, a continuous year term to account for long-term trends, county-month as the conditioning variable, and an offset for county population. In Step 7, we used these models to generate estimates of the relative risk (RR) with 95% confidence intervals (95%CI) of cryptosporidiosis or giardiasis across a matrix of extreme predictor values (i.e., 5^th^, 10^th^, 90^th^ and 95^th^ percentile values) at 2-month lag increments, relative to the risk of disease found at zero (i.e., the mean for a given county and calendar-month).

Finally, to visualize our relative risk estimates and the entire predicted outcome surface across all predictor and lag values, we created a set of plots depicting the predictor-lag-outcome surface, using a reference value of zero (i.e., the mean) in all cases. We used a p-value of <0.05 to indicate statistical significance in this study. Stata 15 (Stata Statistical Software: Release 15 (2017). StataCorp LP, College Station, TX) and R Studio 4.0 (RStudio Team (2020). RStudio: Integrated Development Environment for R. RStudio, PBC, Boston, MA URL http://www.rstudio.com/) were used for all analyses (StataCorp 2015, RStudio Team 2020).

## 3. Results

Between 1997 and 2017, the total population in Colorado increased by more than 40%, rising from approximately 3.9 million to 5.6 million (Table 2). The total count of cryptosporidiosis and giardiasis cases was highest in the summer (38.2% of cryptosporidiosis cases; 30.6% of giardiasis cases) and the fall (32.1% of cryptosporidiosis cases; 30.3% of giardiasis cases) across all Colorado counties. Extreme temperatures (i.e., 5^th^, 10^th^, 90^th^ and 95^th^ percentile values of MC-TMIN and MC-TMAX) were less frequent during the summer season than other times of the year, while precipitation extremes (i.e., 5^th^, 10^th^, 90^th^ and 95^th^ percentile values of MC-PRECIP) were most frequent during the spring and summer seasons (see Table 3). The total Colorado cryptosporidiosis case rate (per 100,000 people) increased over the study period, while Colorado’s giardiasis case rate decreased (Figure 2). There was also a decrease in the frequency of extreme low temperatures (≤10^th^ percentile value of MC-TMIN or MC-TMAX) and a notable increase in the frequency of extreme high temperatures (≥90^th^ percentile value of MC-TMIN or MC-TMAX) in Colorado counties between 1997 and 2017 (Figure 3). See Figure 3 for a depiction of the distribution of Colorado’s county population, county case rates, extreme precipitation, and extreme temperatures in 1997, 2007 and 2017.

**Figure 2.**
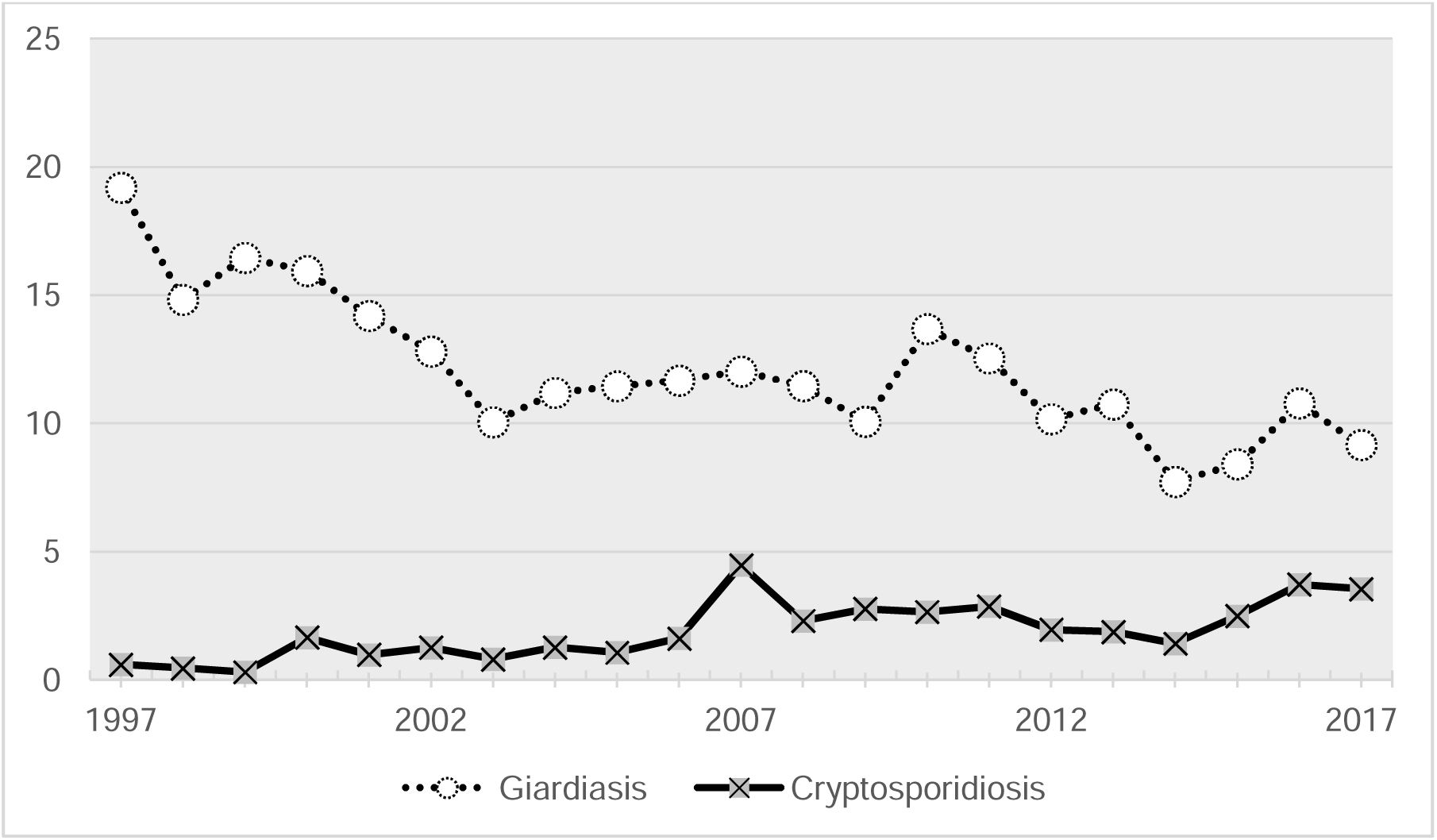
Reported cryptosporidiosis and giardiasis cases across all Colorado counties between 1997 – 2017. **Figure 2 caption:** Depiction of the annual count of cryptosporidiosis and giardiasis cases across all Colorado counties between 1997 – 2017. Whereas there was a decrease in the total number of cases of giardiasis, there was an increase in the total number of cryptosporidiosis cases in Colorado cases between 1997 and 2017.

**Figure 3.**
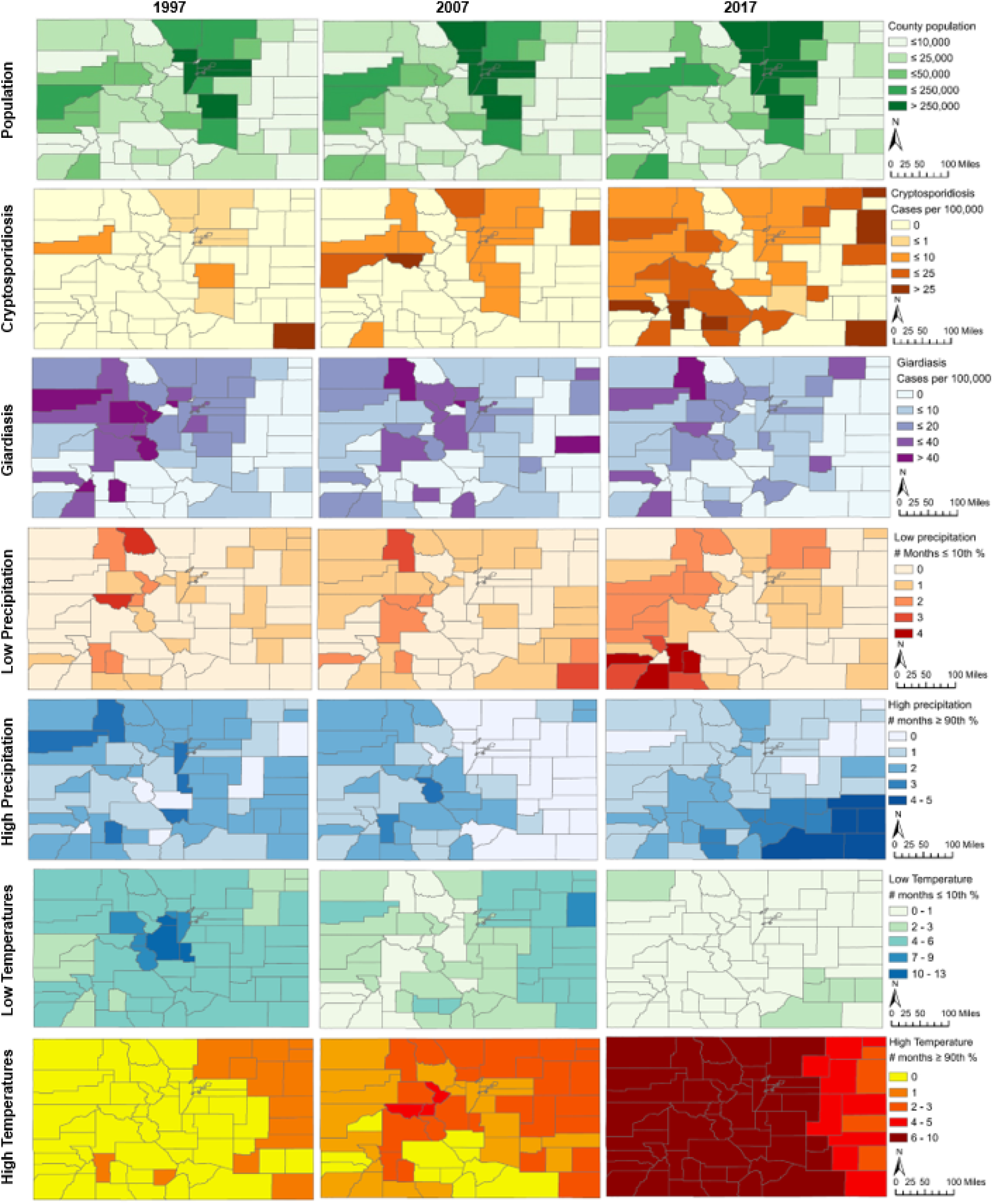
Depiction of population, case rates and extreme weather in 1997, 2007 and 2017 by Colorado counties. **Figure 3 caption:** Maps depicting Colorado county population, cryptosporidiosis and giardiasis case rates (per 100,000 people), and extreme weather in 1997, 2007 and 2017 are shown in Figure 3. The series of maps depicting precipitation indicate the total number of months in 1997, 2007, and 2017 that a county’s mean-centered precipitation was low (≤ 10^th^ percentile value) or high (≥ 90^th^ percentile value), relative to all mean-centered precipitation values observed in Colorado counties across the 21-year study period. The series of maps depicting extreme temperatures combined minimum temperature and maximum temperature, such that the maximum possible number of months with extreme low or extreme high temperature readings for a county in a given year would be 24 (12 possible for maximum temperature, plus 12 possible for minimum temperature). Thus, the low temperature maps indicate the total number of months in 1997, 2007 and 2017 that either the mean-centered minimum temperature or the mean-centered maximum temperature was low (≤ 10^th^ percentile value) for a given county, while the high temperature maps indicate the total number of months in 1997, 2007 and 2017 that either the mean-centered minimum temperature or the mean-centered maximum temperature was high (≥ 90^th^ percentile value) for a given county, relative to all mean-centered minimum and maximum temperature values observed in Colorado counties across the 21-year study period.

**Table 2.** Summary of population and cases at five year increments for Colorado counties during the study period.

| **Summaries by year** | **1997 <sup>a</sup>** | **2002** | **2007** | **2012** | **2017** |
| --- | --- | --- | --- | --- | --- |
| **Population summary** | **N** | **N** | **N** | **N** | **N** |
| Number of counties | 63 <sup>b</sup> | 64 | 64 | 64 | 64 |
| Total population | 3,995,923 | 4,504,709 | 4,821,784 | 5,189,861 | 5,609,445 |
| **Case counts** | **N (% total)** | **N (% total)** | **N (% total)** | **N (% total)** | **N (% total)** |
| Cryptosporidiosis | 24 (1.2) | 57 (2.8) | 216 (10.7) | 102 (5.1) | 199 (9.9) |
| Giardiasis | 767 (6.3) | 577 (4.8) | 580 (4.8) | 528 (4.4) | 514 (4.3) |

<sup>a</sup> The first year that monthly cryptosporidiosis and giardiasis case counts for all Colorado counties became available via the Colorado Electronic Disease Reporting System was 1997. However, our estimates of climatological extremes were generated from data between 1996 – 2017 to allow for lagged effects.
<sup>b</sup> There were 63 counties in Colorado until 2000, when Broomfield county became the 64<sup>th</sup> Colorado county

**Table 3.** Summary of cases and weather extremes by season in Colorado counties between 1996 - 2017.

| Summaries by season | Winter | Spring | Summer | Fall | All Season |
| --- | --- | --- | --- | --- | --- |
| **Case counts** | **N (%)** | **N (%)** | **N (%)** | **N (%)** | **N Total** |
| Cryptosporidiosis | 262 (13.0) | 338 (16.7) | 770 (38.2) | 648 (32.1) | 2,018 |
| Giardiasis | 2,340 (19.4) | 2,386 (19.7) | 3,693 (30.6) | 3,664 (30.3) | 12,083 |
| **Extreme weather summaries <sup>a</sup>** |  |  |  |  |  |
| **MC-TMIN** | **N (%)** | **N (%)** | **N (%)** | **N (%)** | **N Total** |
| ≤ 5<sup>th</sup> percentile (-2.5°C) | 384 (45.5) | 216 (25.6) | 35 (4.2) | 209 (24.8) | 844 |
| ≤ 10<sup>th</sup> percentile (-1.8°C) | 684 (40.5) | 409 (24.2) | 142 (8.4) | 454 (26.9) | 1689 |
| ≥ 90<sup>th</sup> percentile (1.8°C) | 746 (44.2) | 357 (21.1) | 169 (10.0) | 417 (24.7) | 1689 |
| ≥ 95<sup>th</sup> percentile (2.4°C) | 440 (52.1) | 181 (21.4) | 28 (3.3) | 196 (23.2) | 844 |
| **MC-TMAX** | **N (%)** | **N (%)** | **N (%)** | **N (%)** | **N Total** |
| ≤ 5<sup>th</sup> percentile (-3.0°C) | 282 (33.4) | 194 (23.0) | 92 (10.9) | 276 (32.7) | 844 |
| ≤ 10<sup>th</sup> percentile (-2.3°C) | 483 (28.6) | 454 (26.9) | 280 (16.6) | 472 (28.0) | 1689 |
| ≥ 90<sup>th</sup> percentile (2.4°C) | 459 (27.2) | 507 (30.0) | 271 (16.0) | 453 (26.8) | 1689 |
| ≥ 95<sup>th</sup> percentile (3.0°C) | 251 (29.7) | 230 (27.3) | 105 (12.4) | 258 (30.6) | 844 |
| **MC-PRECIP <sup>b</sup>** | **N (%)** | **N (%)** | **N (%)** | **N (%)** | **N Total** |
| ≤ 5<sup>th</sup> percentile (-36.0 mm) | 133 (15.8) | 260 (30.8) | 297 (35.2) | 154 (18.2) | 844 |
| ≤ 10<sup>th</sup> percentile (-28.4 mm) | 232 (13.7) | 530 (31.4) | 554 (32.8) | 373 (22.1) | 1689 |
| ≥ 90<sup>th</sup> percentile (31.5 mm) | 298 (17.6) | 487 (28.8) | 528 (31.3) | 376 (22.3) | 1689 |
| ≥ 95<sup>th</sup> percentile (46.6 mm) | 160 (19.0) | 260 (30.8) | 248 (29.4) | 176 (20.9) | 844 |

MC-TMIN = Countywide monthly average of minimum daily temperature, mean-centered by county-month
MC-TMAX = Countywide monthly average of maximum daily temperature, mean-centered by county-month
MC-PRECIP = Countywide average of the total monthly precipitation, mean-centered by county-month
<sup>a</sup> Extreme temperatures and precipitation were defined as the 5<sup>th</sup>, 10<sup>th</sup>, 90<sup>th</sup> and 95<sup>th</sup> percentile values of the countywide average monthly values of maximum daily temperature, minimum daily temperature and total precipitation, given the county and time of year (i.e., mean-centered by county and month). For each of the four definitions of extreme weather, the total number and percent of all observations that were considered "extreme" using each definition are summarized by three-month season.
<sup>b</sup> Precipitation was summed across all forms of precipitation (e.g. snow, sleet, rain, etc.), and converted to a water-equivalent depth in millimeters.

Each of the final models selected via cross-validation for modeling the impacts of weather extremes on cryptosporidiosis cases used natural or B-splines in both the lag and predictor spaces, while the models for giardiasis cases employed a natural or B-spline in the lag space and a linear function to model the predictor space. Although some of the top six AIC/BIC-selected models included interaction terms to account for some seasonal variation in temperature and precipitation extremes (see Table 3 for details), none of these models were ultimately selected as the final model via cross-validation for any outcome-predictor pair. Appendix Table A3 details the cross-basis structure and the cross-validation results for the top six models of each outcome-predictor pair.

### 3.1. Extreme low temperatures

Compared to the risk of cryptosporidiosis found at the county and calendar-month mean, extreme low temperatures were generally associated with a decrease in cryptosporidiosis risk in the short-term (0 – 8 month lags), followed by an increase in cryptosporidiosis risk in the long-term (10 – 12-month lags) (Figure 4, Table 4). For example, when the maximum temperature was very low (5^th^ percentile) 4-months prior, there was a 6.2% decrease in the risk of cryptosporidiosis cases relative to the risk at the mean (very low MC-TMAX: RR 0.938; 95% CI 0.910 – 0.965), while at the 12-month lag, there was a 16.3% increase (very low MC-TMAX: RR 1.163; 95% CI 1.103 – 1.227). The one exception to the u-shaped pattern between cryptosporidiosis and low temperatures was when minimum temperatures were very low (5^th^ percentile). In this case, our model suggested an inverted u-shaped wherein there was an increase in cryptosporidiosis risk (relative to the risk at the county and calendar-month mean) at all lags between 0 and 12-months, which peaked between the 6-month lag (very low MC-TMIN: RR 1.080, 95%CI 1.046 – 1.116) and 8-month lag (very low MC-TMIN: RR: 1.083, 95% CI 1.052 – 1.114).

**Figure 4.**
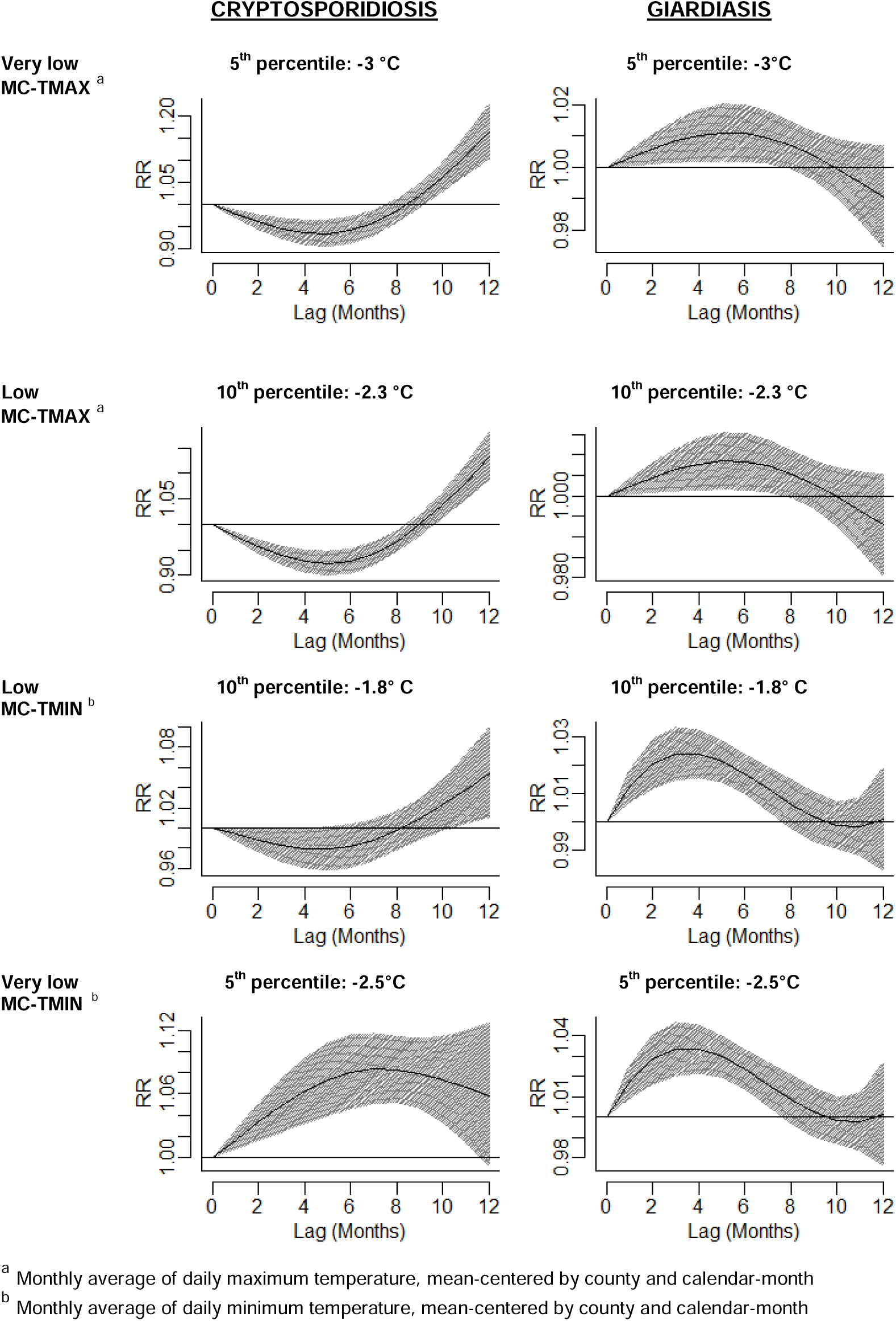
Depictions of the lagged effects of extreme low temperatures on the risk of cryptosporidiosis and giardiasis cases in Colorado counties. **Figure 4 caption:** The effects of very low (5^th^ percentile) and low (10^th^ percentile) temperatures, lagged 0 – 12 months, on the risk of cryptosporidiosis and giardiasis (relative to the risk at the county and calendar month mean) are depicted in Figure 3. The top panel depicts extreme low values of the monthly average daily maximum temperature, mean-centered by county and calendar-month (MC-TMAX), while the bottom panel depicts extreme low values of the monthly average daily minimum temperature, mean-centered by county and calendar-month (MC-TMIN).

**Table 4.**
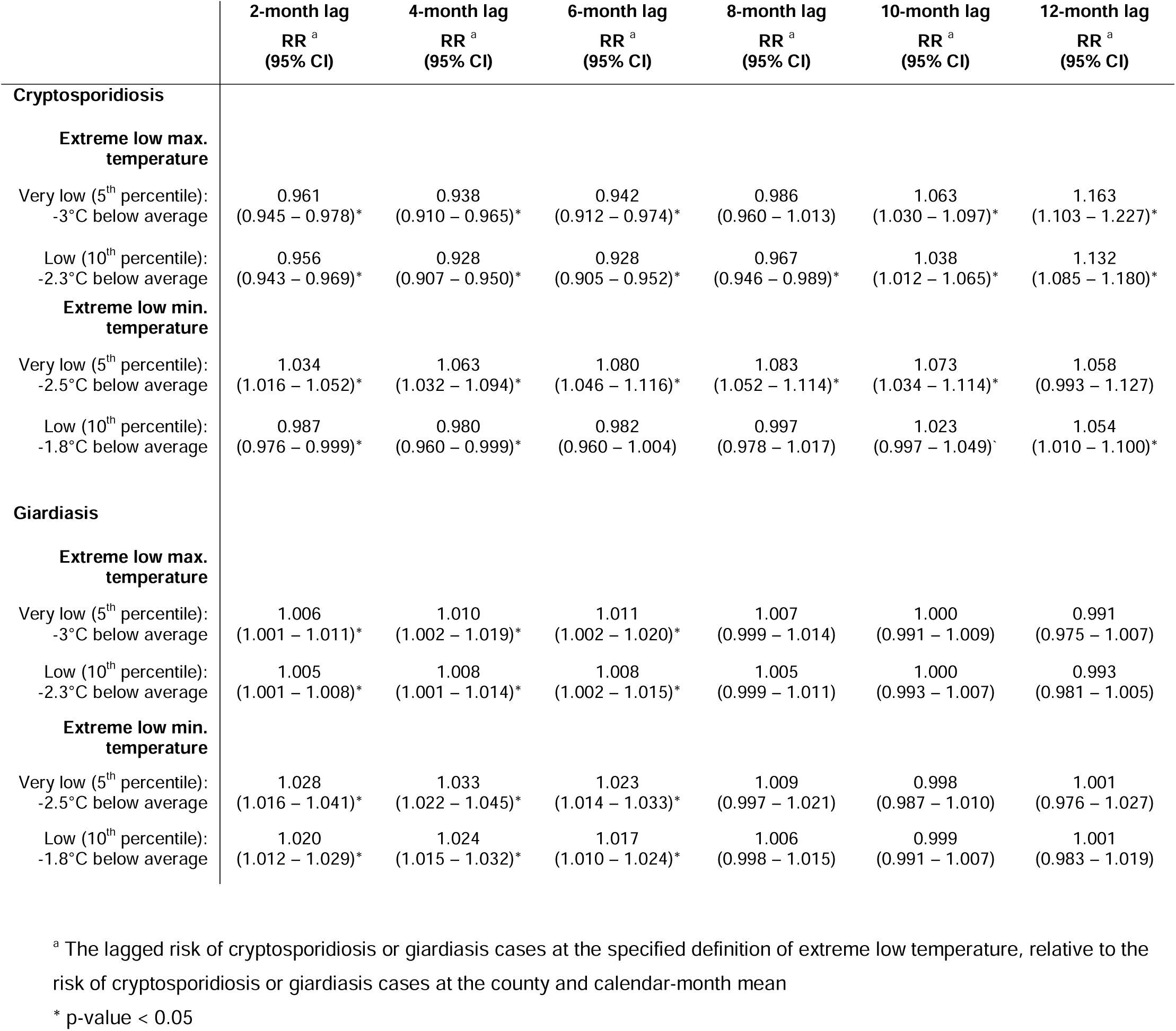
Comparing the lagged effects of extreme low temperatures on the relative risk of cryptosporidiosis and giardiasis cases in Colorado counties between 1997 – 2017.

We also found an inverted u-shaped pattern between lagged extreme low temperatures and giardiasis (Figure 4, Table 4). When maximum temperatures were very low (5^th^ percentile) or low (10^th^ percentile), the relative risk of giardiasis cases relative to the risk at the county and calendar-month mean was highest at the 6-month lag (very low MC-TMAX: RR 1.011, 95% CI 1.002 – 1.020; low MC-TMAX: RR 1.008, 95% CI 1.002 – 1.015). By comparison, the relative risk of giardiasis peaked at the 4-month lag for both measures of minimum temperatures (very low MC-TMIN: RR 1.033; 95% CI 1.022 – 1.045; low MC-TMIN: RR 1.024; 95% CI 1.015 – 1.032). At longer lags (8 – 12 months), we found that neither low nor very low values of MC-TMAX or MC-TMIN yielded a significant change in the risk of giardiasis cases, relative to the mean.

### 3.2. Extreme high temperatures

When MC-TMAX or MC-TMIN was either high (90^th^ percentile) or very high (95^th^ percentile), there was an inverted u-shaped increase in the lagged (0 to 12-months) risk of cryptosporidiosis, relative to the risk at the county and calendar-month mean (Figure 5). In all cases, the relative risk of cryptosporidiosis peaked at the 6-month lag (high MC-TMAX: RR 1.086; 95% CI 1.064 – 1.109; very high MC-TMAX: RR 1.076; 95% CI 1.048 – 1.105; high MC-TMIN: RR 1.129; 95% CI 1.104 – 1.155; very high MC-TMIN: RR 1.122, 95% CI 1.087 – 1.157) (Table 5).

**Figure 5.**
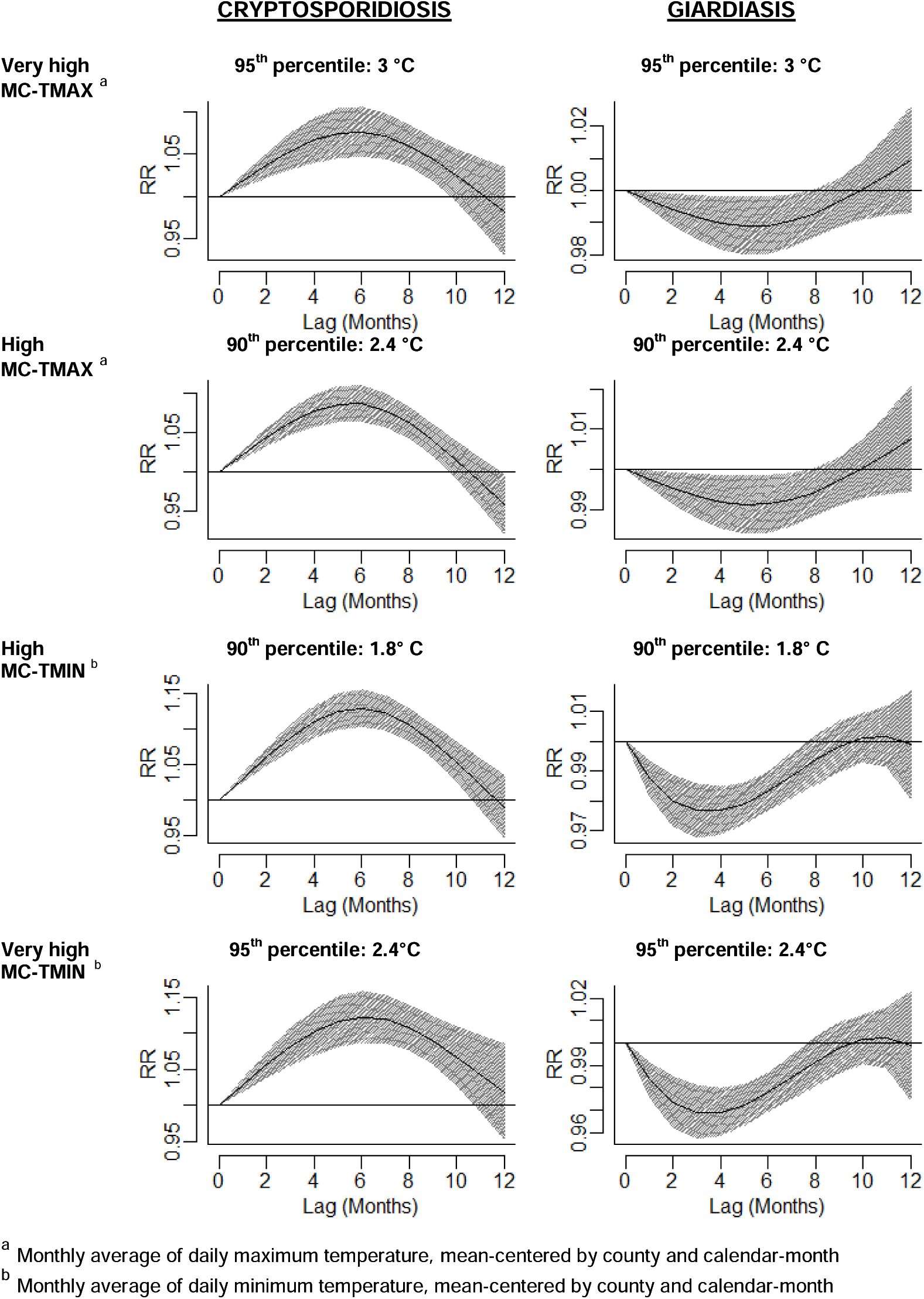
Depictions of the lagged effects of extreme high temperatures on the risk of cryptosporidiosis and giardiasis cases in Colorado counties. **Figure 5 caption:** The effects of very high (95^th^ percentile) and high (90^th^ percentile) temperatures, lagged 0 – 12 months, on the risk of cryptosporidiosis and giardiasis (relative to the risk at the county and calendar month mean) are depicted in Figure 4. The top panel depicts extreme high values of the monthly average daily maximum temperature, mean-centered by county and calendar-month (MC-TMAX), while the bottom panel depicts extreme high values of the monthly average daily minimum temperature, mean-centered by county and calendar-month (MC-TMIN).

**Table 5.**
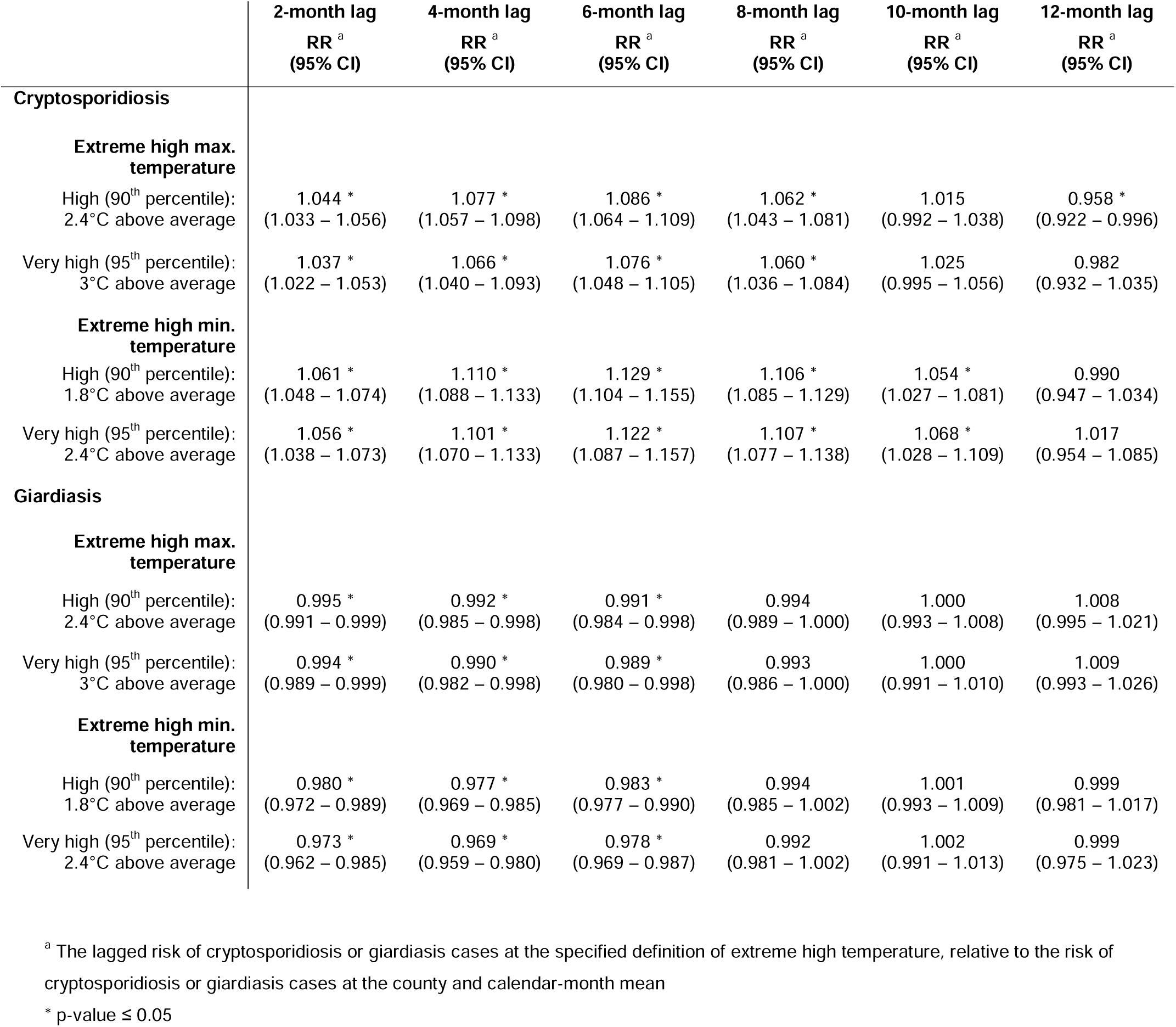
Comparing the lagged effects of extreme high temperatures on the relative risk of cryptosporidiosis and giardiasis cases in Colorado counties between 1997 – 2017.

For giardiasis, we found a significant decrease in the risk of cases when MC-MAXT or MC-MINT were lagged between 0 – 6 months and either high (90^th^ percentile) or very high (95^th^ percentile) (Figure 4). The risk of giardiasis at high or very high temperatures (relative to the risk at the county and calendar month mean values) was at its lowest at the 4-month lag in all cases (high MC-TMAX: RR 0.991; 95% CI 0.984 – 0.998; very high MC-TMAX: RR 0.989; 95% CI 0.980 – 0.998; high MC-TMIN: RR 0.983; 95% CI 0.977 – 0.990; very high MC-TMIN: RR 0.978, 95% CI 0.969 – 0.987) (Table 5). The decrease in the relative risk of giardiasis was more pronounced with extreme high minimum temperatures than with extreme high maximum temperatures, as shown in Figure 5.

### 3.3. Extreme low precipitation

The relationship between extreme low levels of monthly precipitation and both cryptosporidiosis and giardiasis risk was only significantly different than the risk at the county and calendar-month mean when allowing for a lag of at least 8-months (Figure 6). In all cases, our models suggested that the relative risk of both cryptosporidiosis and giardiasis was lowest 12-months after low (10^th^ percentile) or very low (5^th^ percentile) levels of precipitation (Table 6). In the case of cryptosporidiosis, there was an 18.1% decrease (RR 0.819, 95% CI 0.769 – 0.871) in the risk of cases relative to the county and calendar-month mean when precipitation 12-months prior was very low (5^th^ percentile), and a 13.5% decrease (RR 0.865, 95% CI 0.825 – 0.905) in the risk of cases when precipitation 12-months prior was low (10^th^ percentile). For giardiasis, the associations with extreme low precipitation were similar, but less pronounced: there was a 3.2 decrease (RR 0.977, 95% CI 0.962 – 0.992) in the relative risk of giardiasis cases when precipitation 12-months prior was very low (5^th^ percentile), and a 1.8% decrease (RR 0.982, 95% CI 0.970 – 0.994) in the risk of cases when precipitation 12-months prior was low (10^th^ percentile).

**Figure 6.**
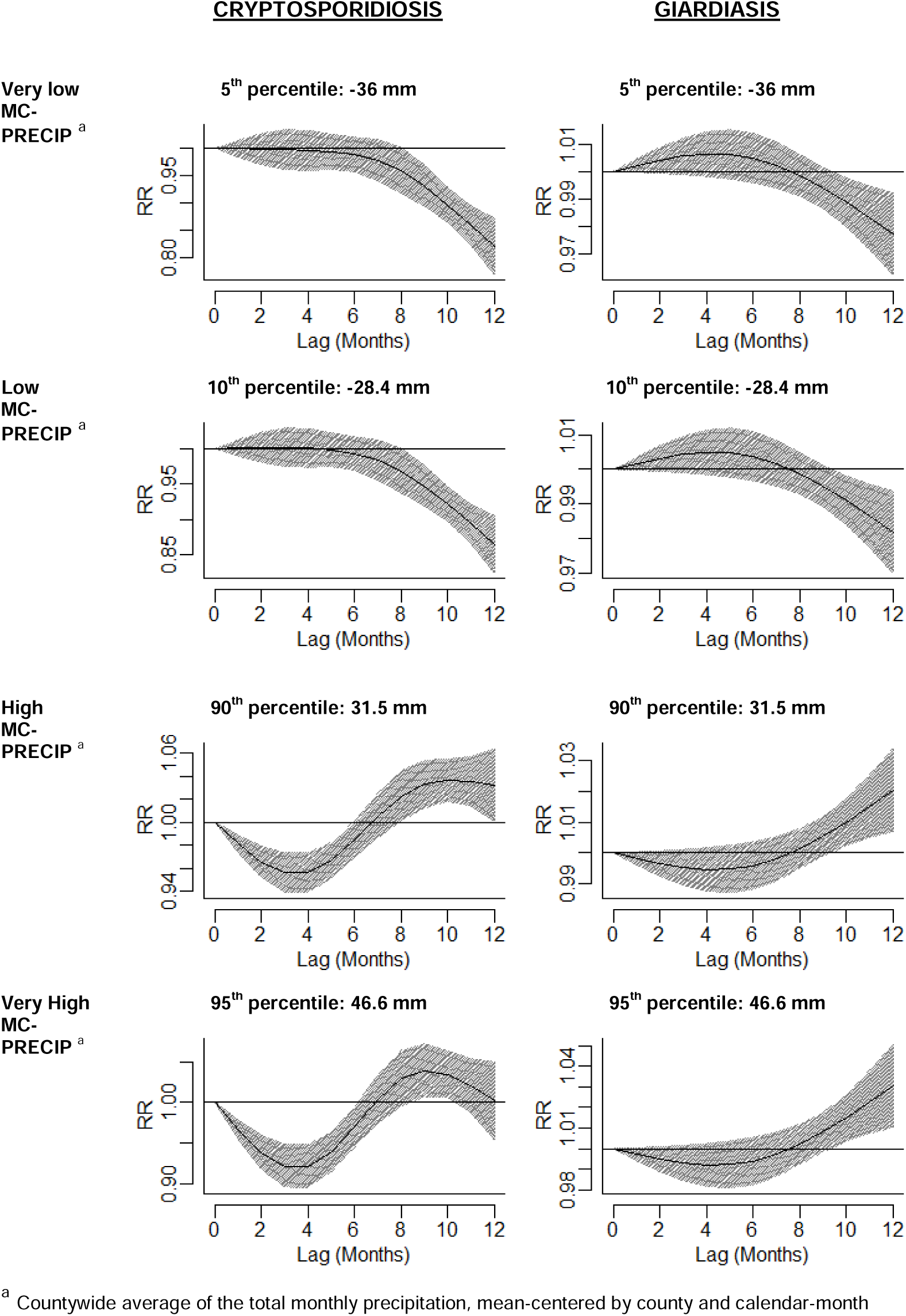
Depictions of the lagged effects of extreme precipitation on the risk of cryptosporidiosis and giardiasis cases in Colorado counties. **Figure 6 caption:** The effects of extreme monthly precipitation, lagged 0 – 12 months, on the risk (relative to the risk at the county and calendar-month mean) of cryptosporidiosis (left) and giardiasis (right) are depicted in Figure 5. The top panel depicts the lagged effects of very low (5^th^ percentile) and low (10^th^ percentile) values of the countywide average of the total monthly precipitation, mean-centered by county and calendar-month (MC-PRECIP), while the bottom panel depicts the lagged effects of very high (95^th^ percentile) and high (90^th^ percentile) values of MC-PRECIP on the risk of cryptosporidiosis and giardiasis.).

**Table 6.**
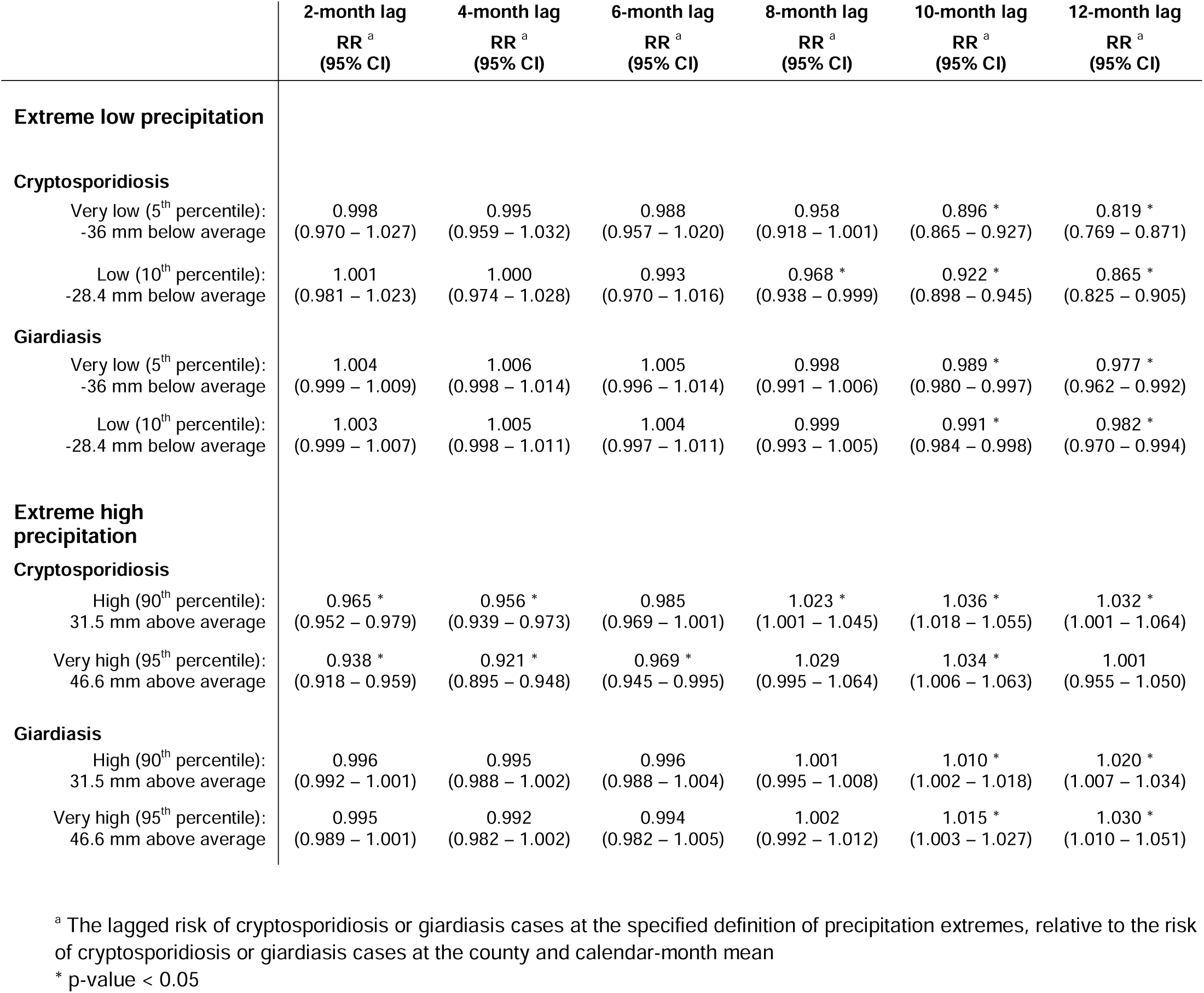
Comparing the lagged effects of precipitation extremes on the relative risk of cryptosporidiosis and giardiasis cases in Colorado counties between 1997 – 2017.

|  | 2-month lag<br>RR<sup>a</sup><br>(95% CI) | 4-month lag<br>RR<sup>a</sup><br>(95% CI) | 6-month lag<br>RR<sup>a</sup><br>(95% CI) | 8-month lag<br>RR<sup>a</sup><br>(95% CI) | 10-month lag<br>RR<sup>a</sup><br>(95% CI) | 12-month lag<br>RR<sup>a</sup><br>(95% CI) |
| --- | --- | --- | --- | --- | --- | --- |
| **Extreme low precipitation** |  |  |  |  |  |  |
| **Cryptosporidiosis** |  |  |  |  |  |  |
| Very low (5<sup>th</sup> percentile):<br>-36 mm below average | 0.998<br>(0.970 – 1.027) | 0.995<br>(0.959 – 1.032) | 0.988<br>(0.957 – 1.020) | 0.958<br>(0.918 – 1.001) | 0.896 *<br>(0.865 – 0.927) | 0.819 *<br>(0.769 – 0.871) |
| Low (10<sup>th</sup> percentile):<br>-28.4 mm below average | 1.001<br>(0.981 – 1.023) | 1.000<br>(0.974 – 1.028) | 0.993<br>(0.970 – 1.016) | 0.968 *<br>(0.938 – 0.999) | 0.922 *<br>(0.898 – 0.945) | 0.865 *<br>(0.825 – 0.905) |
| **Giardiasis** |  |  |  |  |  |  |
| Very low (5<sup>th</sup> percentile):<br>-36 mm below average | 1.004<br>(0.999 – 1.009) | 1.006<br>(0.998 – 1.014) | 1.005<br>(0.996 – 1.014) | 0.998<br>(0.991 – 1.006) | 0.989 *<br>(0.980 – 0.997) | 0.977 *<br>(0.962 – 0.992) |
| Low (10<sup>th</sup> percentile):<br>-28.4 mm below average | 1.003<br>(0.999 – 1.007) | 1.005<br>(0.998 – 1.011) | 1.004<br>(0.997 – 1.011) | 0.999<br>(0.993 – 1.005) | 0.991 *<br>(0.984 – 0.998) | 0.982 *<br>(0.970 – 0.994) |
| **Extreme high precipitation** |  |  |  |  |  |  |
| **Cryptosporidiosis** |  |  |  |  |  |  |
| High (90<sup>th</sup> percentile):<br>31.5 mm above average | 0.965 *<br>(0.952 – 0.979) | 0.956 *<br>(0.939 – 0.973) | 0.985<br>(0.969 – 1.001) | 1.023 *<br>(1.001 – 1.045) | 1.036 *<br>(1.018 – 1.055) | 1.032 *<br>(1.001 – 1.064) |
| Very high (95<sup>th</sup> percentile):<br>46.6 mm above average | 0.938 *<br>(0.918 – 0.959) | 0.921 *<br>(0.895 – 0.948) | 0.969 *<br>(0.945 – 0.995) | 1.029<br>(0.995 – 1.064) | 1.034 *<br>(1.006 – 1.063) | 1.001<br>(0.955 – 1.050) |
| **Giardiasis** |  |  |  |  |  |  |
| High (90<sup>th</sup> percentile):<br>31.5 mm above average | 0.996<br>(0.992 – 1.001) | 0.995<br>(0.988 – 1.002) | 0.996<br>(0.988 – 1.004) | 1.001<br>(0.995 – 1.008) | 1.010 *<br>(1.002 – 1.018) | 1.020 *<br>(1.007 – 1.034) |
| Very high (95<sup>th</sup> percentile):<br>46.6 mm above average | 0.995<br>(0.989 – 1.001) | 0.992<br>(0.982 – 1.002) | 0.994<br>(0.982 – 1.005) | 1.002<br>(0.992 – 1.012) | 1.015 *<br>(1.003 – 1.027) | 1.030 *<br>(1.010 – 1.051) |

<sup>a</sup> The lagged risk of cryptosporidiosis or giardiasis cases at the specified definition of precipitation extremes, relative to the risk of cryptosporidiosis or giardiasis cases at the county and calendar-month mean
\* p-value < 0.05

### 3.4. Extreme high precipitation

When precipitation was high (90^th^ percentile) or very high (95^th^ percentile), the risk of cryptosporidiosis cases (relative to the risk at the county and calendar-month mean) decreased in the short-term (2 – 6 month lags), but increased relative to the mean at lags of 8 – 12 months (see Figure 6).

In the short-term, the decrease in cryptosporidiosis risk was largest 4-months after higher than average precipitation (high MC-PRECIP: RR 0.956, 95% CI 0.939 – 0.973; very high MC-PRECIP: RR 0.921, 95% CI 0.895 – 0.948) while in the long-term, the relative increase in cryptosporidiosis risk peaked at 10-months post high precipitation levels (high MC-PRECIP: RR 1.036, 95% CI 1.018 –1.055; very high MC-PRECIP: RR 1.034, 95% CI 1.006 – 1.063).

In the case of giardiasis, extreme high precipitation was not associated with a significant change in giardiasis risk in the short-term (relative to the risk at county and calendar-month mean precipitation levels), but was associated with a small increase in risk in the long-term (i.e., 10 and 12-month lags) (Figure 6, Table 6). For example, when precipitation was 31.5 mm above the monthly average (90^th^ percentile) 12-months prior, there was a 2% increase in the risk of giardiasis cases (RR 1.020, 95% CI 1.007 – 1.034), while at 46.6 mm above the monthly average (5^th^ percentile) 12-months prior, there was a 3% increase in the risk of giardiasis cases (RR 1.030, 95% CI 1.010 – 1.051).

## 4. Discussion

In this investigation of the relationships between weather extremes and two highly burdensome parasitic diseases in Colorado counties between 1997 and 2017, we found distinctly different patterns in the associations between temperature extremes and cryptosporidiosis, versus temperature extremes and giardiasis. Conversely, our models of precipitation extremes were very similar between cryptosporidiosis and giardiasis, all of which highlighted the prominent role of long-term lags. These are novel findings that have not been replicated in other epidemiological studies, which may, at least in part, be due to the unique methodology employed in the current study. Whereas other studies to date have tended to focus on short-lived extreme weather events that represent relatively infrequent occurrences (e.g., flooding), or simple, absolute definitions of extremes (e.g., ≥ 37 °C) and lags (e.g., 3-months), in our study, extremes were always defined relative to the county and calendar-month mean, and lags were more flexible as a result of our use of DLNMs. As such, our analysis provides novel insights into the subtleties of the lagged relationships between periodic temperature and precipitation extremes relative to local averages and two important, but often overlooked protozoan diseases over an extended period.

In the case of extreme low temperatures, we found a significant increase in giardiasis case risk when either the maximum or minimum temperature (lagged ≤ 6 months) was low or very low (10^th^ and 5^th^ percentile, respectively) relative to the county and calendar-month. By contrast, we found a significant decrease in the relative risk of cryptosporidiosis when maximum temperature was low or very low 2-8 months prior, as well when minimum temperature was low 2-4 months prior (though notably, the relationship was reversed when minimum temperature was very low). To date, few studies have focused on the relationship between extreme low temperatures and cryptosporidiosis or giardiasis, and those that have tend to be experimental and focused on parasite viability and survival rather than real-world transmission dynamics and risks. This dearth of epidemiological evidence regarding protozoa and low temperatures may be linked to a historical focus on transmission potential at above average temperatures, as investigations into the seasonality of giardiasis and cryptosporidiosis have established that incidence tends to peak in spring or summer in temperate climates (Naumova, Jagai et al. 2007, Jagai, Castronovo et al. 2009, Lal, Hales et al. 2012, Gutiérrez-Gutiérrez and Palomo-Ligas 2023).

The relationship that we found between extreme high temperatures and cryptosporidiosis and giardiasis risk was consistent with that of low temperatures: when maximum or minimum temperatures were high (90^th^ percentile) or very high (95^th^ percentile), we found a significant increase in cryptosporidiosis risk, but a significant decrease in giardiasis risk, relative to the risk found at the county and calendar-month mean. For both cryptosporidiosis and giardiasis, the relative risk peaked 6-months after extreme high temperatures, though for cryptosporidiosis, there remained a statistically significant association at longer lags (i.e., 8 and 10 months). In the case of cryptosporidiosis, our findings align with the existing evidence on the relationship with high maximum/minimum temperatures, which suggests that there tends to be higher risk of cryptosporidiosis at higher temperatures (Ajjampur, Liakath et al. 2010, Hu, Mengersen et al. 2010, Kent, McPherson et al. 2015, Brankston, Boughen et al. 2018). For example, one study conducted in Brisbane Australia demonstrated that when relative humidity was less than 63% and weekly maximum temperature exceeded 31°C, the relative risk of cryptosporidiosis cases increased by 13.64 (Hu, Mengersen et al. 2010). While research on giardiasis and extreme temperature is sparse, studies on *Giardia* cyst survival and identification also align with our findings, suggesting a negative association between air and water temperatures and concentrations of *Giardia* cysts in the ambient environment (Wilkes, Edge et al. 2011, Li, Chase et al. 2019, Masina, Shirley et al. 2019). However, it is worth noting that these studies also found that the same was true of *Cryptosporidium* oocysts (Wilkes, Edge et al. 2011, Li, Chase et al. 2019, Masina, Shirley et al. 2019), which conflicts with the aforementioned studies suggesting that cryptosporidiosis risk increases with higher temperatures (Ajjampur, Liakath et al. 2010, Hu, Mengersen et al. 2010, Kent, McPherson et al. 2015, Brankston, Boughen et al. 2018). Taken together, these seemingly contradictory results may suggest that both extreme low and extreme high temperatures have the potential to amplify cryptosporidiosis risk under the right environmental conditions, whereas *Giardia* may simply be more sensitive to extreme high temperatures. This is supported by evidence from in vitro and in vivo experiments which have indicated that *Giardia* cysts demonstrate greater temperature-dependence and higher die-off rates than *Cryptosporidium* oocysts at both high and low temperatures (Olson, Goh et al. 1999, Alum, Absar et al. 2014).

Another possible explanation for why *Cryptosporidium* and *Giardia* respond differently to extreme temperatures is that the primary hosts associated with each pathogen are different. For example, a study conducted in Georgia, USA found that synanthropic flies living alongside wildlife and livestock could serve as mechanical vectors of *Cryptosporidium* and *Giardia*, with viable *Cryptosporidium* being isolated from 56% of sampled flies, while *Giardia* was isolated from just 8% of those sampled (Conn, Weaver et al. 2007). As warm-weather, ectothermic species, the development time of the studied fly species tends to decrease as temperatures increase, up until a threshold temperature when development will cease – for example, above 34°C for similar species of blowflies found in Asia (Zhang, Wang et al. 2019). Thus, if cattle and synanthropic flies are in fact a major part of the transmission pathway for cryptosporidiosis, but not giardiasis in Colorado, then the association between cryptosporidiosis (and not giardiasis) and above average maximum and minimum temperatures (which notably, generally fall below a 34°C threshold in Colorado counties) is consistent with what would be expected. While further study is still needed to determine whether flies are in fact an important vector in either the cryptosporidiosis or giardiasis transmission cycles in Colorado, epidemiological studies to date have highlighted that livestock (primarily cattle) are a major reservoir of zoonotic *Cryptosporidium parvum*, and that direct contact with livestock or indirect contact via drinking water are likely major transmission pathways (Fayer, Morgan et al. 2000, Stantic-Pavlinic, Xiao et al. 2003, Hunter and Thompson 2005). Meanwhile, livestock are not believed to be a major source of giardiasis transmission to human populations (Hunter and Thompson 2005).

In this study, we found that extreme high levels of precipitation were followed by a decrease in the relative risk of cryptosporidiosis (and giardiasis, though not statistically significant) in the short-term (<8-month lag), and a significant increase in the relative risk of both cryptosporidiosis and giardiasis in the long-term (10 – 12 month lags). Although the lags that we used in our study were considerably longer than the typical lags assessed for cryptosporidiosis and giardiasis to date, these findings may potentially be indicative of a dilution effect in the short-term, followed by increased dispersal and opportunity for transmission events in the long-term. While evidence in favor of the latter phenomenon is difficult to assess given that long-term risks of extreme precipitation have not been a focus in the literature to date, one study from Australia did find a 1.8% increase in cryptosporidiosis risk when annual rainfall increased by 78 mm (Forbes, Hosking et al. 2021). Meanwhile, several studies have provided evidence in favor of a short-term dilution effect. For example, the dilution effect was believed to be at play in one study conducted in Canada, which demonstrated lower odds of human cryptosporidiosis when water levels were higher than average 19-20 days prior (Brankston, Boughen et al. 2018). Similarly, an assessment of environmental concentrations of giardia cysts in storm-based runoff from dairy lots in California found an overall negative association between giardia concentrations and cumulative precipitation, with the highest concentrations found early in the rainfall-runoff season (Miller, Lewis et al. 2007). Notably, a recent review that assessed the body of evidence in support of the concentration-dilution hypothesis – that is, that conflicting findings on the impacts of rainfall on diarrhea result from underlying differences in background rain levels – found four studies that identified dilution (specifically, rainfall following wet periods) as a potential mechanism explaining an inverse association between rainfall and diarrhea (Kraay, Man et al. 2020). While an investigation of the impacts of rainfall following wetter than average weather was outside of the scope of this study, future investigations of the impacts of this phenomenon on giardiasis and cryptosporidiosis, as well as the underlying sources of contamination for each pathogen within Colorado counties is warranted.

In the case of extreme low levels of precipitation, we found that netiher low nor very low precipitation was significantly associated with the relative risk of either cryptosporidiosis or giardiasis cases in the short-term (<8-month lag), but in the long-term (10 – 12 month lags), there was a significant decrease in the risk of both cryptosporidiosis and giardiasis cases. Whereas other studies assessing the effects of weather and climate on cryptosporidiosis and giardiasis have most frequently assessed lag periods of between one and three months (Hu, Tong et al. 2007, Naumova, Jagai et al. 2007, Hu, Mengersen et al. 2010, Hu, Mengersen et al. 2010, Kent, McPherson et al. 2015), in our study, we allowed for a lag of up to 12-months. This decision was based on the body of evidence from a small set of experimental studies which have suggested that under the right conditions, *Cryptosporidium* oocysts and *Giardia* cysts may be able to survive and maintain viability for much longer than the period typically assessed in the epidemiological literature – potentially as long as six months to a year (Badenoch 1990, Fayer, Trout et al. 1998). Overall, our analysis suggest that that the transmission of cryptosporidiosis and giardiasis in Colorado counties may be sensitive to longer-term periods of excess dryness or drought, as has been found with diarrheal pathogens in other regions of the world (Bush, O’Neill et al. 2014, Chhetri, Takaro et al. 2017, Lal and Konings 2018). While our study was not designed to assess the impacts of rainfall after prolonged periods of drought on transmission, future studies aimed at investigating this in Colorado and the American West are greatly needed.

Although giardiasis and cryptosporidiosis cases reported in Colorado counties between 1997 – 2017 tended to follow the standard seasonal pattern that have been previously identified for temperate climates, wherein case counts tend to peak in the summer or early fall (Lal, Hales et al. 2012) (See Table 3), the final models selected via cross-validation for each of our outcome-predictor pairs did not ultimately include an interaction for season. This is notable, as season-specific effects have been demonstrated in other contexts (Odoi, Martin et al. 2003, Muchiri, Ascolillo et al. 2009), which are believed to arise as a result of season-specific exposure opportunities such as local agricultural practices, recreational water uses, and how people interact with the environment (Learmonth, Ionas et al. 2002, Odoi, Martin et al. 2003). While it is possible that the relationship between precipitation and temperature and cryptosporidiosis and giardiasis truly does not vary substantially by season in Colorado counties, it is also possible that our models were underpowered for detecting seasonal effects.

Thus, a limitation of this study was the potential for being underpowered when it came to the seasonal effect modification assessment, due to the panel structure of the data, our conditioning on county-month, the complex lag-predictor cross-basis structures already included in each model, and the fact that we had a relatively small number of observations for each county (252 month-years for 63 counties, and 216 for Broomfield County). Another limitation to this study was that the smallest time-unit possible given the data available for the study period was month, making it impossible for us to determine the effects of short-term weather events. Additionally, because county was the smallest geographic-unit in this study, we were unable to investigate sub-county location-specific nuances such as variations in elevation, access to healthcare (which may impact case reporting) or popular recreational activities (which may impact exposure). While including a grouping term in our cross-basis and conditioning on county-calendar-month helps to account for unmeasured intra-county variation, we remain unable to assess inner-county differences.

Another limitation to this study is that our outcome data does not necessarily indicate the location where the pathogen was acquired, only the county of residence for an infected individual. This means that travel related cases (i.e., cases acquired in another county, state or country) were not excluded from our analysis, and were therefore a probable source of noise in our data that likely biased our results toward the null. This study also relied on laboratory reported cryptosporidiosis and giardiasis, and, due to the variable severity of disease, many infections with these protozoa go unreported. Meanwhile, among cases of sufficient severity to warrant medical attention, changes in the use, accuracy and types of diagnostic tests that were available for the clinical detection of cryptosporidiosis and giardiasis across the study period also have the potential to bias our results. While cryptosporidiosis and giardiasis diagnostic tests remained relatively unchanged in the 90s and early 2000s (Ricciardi and Ndao 2015), the more widespread introduction of multiplex tests means that healthcare providers could more easily test for an array of diseases in patients exhibiting gastrointestinal symptoms. Being able to easily test for multiple diseases could increase detection of suspected disease, as well as increase incidental findings of cryptosporidiosis and giardiasis. However, as shown in Figure 2, giardiasis cases have decreased in Colorado over time, whereas cryptosporidiosis cases have steadily increased. A linear term for year was included in each of our models to help account for these and other linear time trends.

## 5. Conclusion

Our analysis has shown that the risk of cryptosporidiosis and giardiasis in Colorado and beyond is likely to change as weather patterns shift in the coming years. Warmer temperatures and an increase in weather extremes can be expected. In the last 30 years, Colorado’s average temperature has increased by 2°F (1.1 °C), with current estimates projecting that the average temperature for the state could increase another 5°F (2.8°C) before 2050 (Colorado Health Institute 2017). As such, it is possible that the coming years will see a continued reduction in the transmission of giardiasis (Figure 2), as our study has highlighted that extreme high temperatures tend to be associated with reduced giardiasis infection risk in Colorado. Meanwhile, our results have also suggested the potential for increasing cryptosporidiosis risk in the years to come, as we found extreme high temperatures were associated with an increase in the risk of cryptosporidiosis cases. These discordant findings indicate that the impact of climate change may not be the same for all protozoan pathogens and highlight the need for further study of the location-specific environmental conditions that could be simultaneously promoting the transmission potential of one protozoan pathogen while suppressing another. Our study has also highlighted the potential for long-term lagged effects of extreme precipitation on cryptosporidiosis and giardiasis, a novel finding that warrants further investigation in other contexts. As the frequency and magnitude of temperature and precipitation extremes rise as a result of human-driven climate change, climate extremes are gradually shifting to become the “new normal”. As such, our analysis contributes to the larger body of science on climate and health, helping to characterize the diverse and complex effects that our changing climate is likely having on human health now and in the not-so-distant future.

## Supporting information

AppendixTables1-3

## CRediT authorship contribution statement

**Elise Grover:** Conceptualization, Methodology, Software, Validation, Formal analysis, Data curation, Writing – Original Draft, Writing – Review & Editing, Visualization, Project administration. **James Crooks:** Methodology, Software, Validation, Writing – Original Draft, Writing – Review & Editing, Supervision. **Elizabeth Carlton**: Writing – Original Draft, Writing – Review & Editing, Supervision. **Sara Paull:** Writing – Original Draft, Writing – Review & Editing. **William Allshouse:** Writing – Original Draft, Writing – Review & Editing. **Rachel Jervis:** Resources, Data Curation, Writing – Review & Editing. **Katherine James:** Conceptualization, Methodology, Data curation, Writing – Original Draft, Writing – Review & Editing, Supervision, Project administration, Funding acquistion.

## Acknowledgements

This work was made possible by the Colorado Department of Public Health and Environment (CDHPE) and the National Center for Atmospheric Research (NCAR), who collected and shared the data used in this analysis.

## Data availability

Data on reportable diseases in Colorado is made available upon request from CDPHE. The Daily Surface Weather and Climatological Summaries (Daymet) Version 3 dataset that was used in this analysis is available upon request from NCAR.

