## AppendixTables1-3 for "Investigating the relationship between extreme weather and cryptosporidiosis and giardiasis in Colorado: a multi-decade study using distributed-lag nonlinear models"

**Appendix Table A1. Defining extreme temperature and precipitation relative to the mean for the time of year among Colorado counties.**

| **Key values across predictor distribution** | **Difference in mean-centered minimum monthly temperature (MC-TMIN) (°C)** | **Difference in mean-centered maximum monthly temperature (MC-TMAX) (°C)** | **Difference in mean-centered total monthly precipitation (MC-PRECIP) (mm)** | **Qualitative definition** |
| --- | --- | --- | --- | --- |
| 5^th^ percentile | -2.5°C | -3.0°C | -36.0 mm | Very low, relative to the calendar-month and county mean |
| 10^th^ percentile | -1.8°C | -2.3°C | -28.4 mm | Low, relative to the calendar-month and county mean |
| Mean | 0°C | 0 °C | 0 mm | Calendar-month and county mean |
| 90^th^ percentile | 1.8°C | 2.4°C | 31.5 mm | High, relative to the calendar-month and county mean |
| 95^th^ percentile | 2.4°C | 3.0°C | 46.6 mm | Very high, relative to the calendar-month and county mean |

To assess the impacts of “extreme” weather relative to “normal” weather on cryptosporidiosis and giardiasis case counts in Colorado counties between 1997 - 2017, TMIN, TMAX and PRECIP were all mean-centered (MC) by county and calendar-month. The 5^th^, 10^th^, 90^th^ and 95^th^ percentile values of MC-TMIN, MC-TMAX, and MC-PRECIP across all counties and months are detailed above. For the purposes of this analysis, the 5^th^ and 95^th^ are referred to as “very” low/high values, relative to the calendar-month and county mean, and the 10^th^ and the 90^th^ are referred to as “low” and “high”, relative to the calendar-month and county mean.

**Appendix Table A2. Model selection for assessing the impacts of minimum monthly temperature on cryptosporidiosis.**

| **Model Name** | **Predictor function** | **Predictor DF / Knot** | **Lag function** | **Lag DF** | **Season**  **Interaction** | **BIC** | **AIC** |
| --- | --- | --- | --- | --- | --- | --- | --- |
| **Step 2: Lag space model variants** |  |  |  |  |  |  |  |
| Crypt_TMIN_Lin_NS2 | Linear | NA | N-spline | 2 | No | 15419.9*** | 9493.9 |
| Crypt_TMIN_Lin_NS3 | Linear | NA | N-spline | 3 | No | 15427.9 | 9494.2 |
| Crypt_TMIN_Lin_NS4 | Linear | NA | N-spline | 4 | No | 15430.3 | 9489.0 |
| Crypt_TMIN_Lin_NS5 | Linear | NA | N-spline | 5 | No | 15436.1 | 9487.1 |
| Crypt_TMIN_Lin_NS6 | Linear | NA | N-spline | 6 | No | 15442.2 | 9485.5 |
| Crypt_TMIN_Lin_NS7 | Linear | NA | N-spline | 7 | No | 15452.3 | 9487.2 |
| Crypt_TMIN_Lin_PS5 | Linear | NA | P-spline | 5 | No | 15436.2 | 9487.2 |
| Crypt_TMIN_Lin_PS6 | Linear | NA | P-spline | 6 | No | 15446.7 | 9490.0 |
| Crypt_TMIN_Lin_PS7 | Linear | NA | P-spline | 7 | No | 15455.6 | 9491.2 |
| Crypt_TMIN_Lin_BS3 | Linear | NA | B-spline | 3 | No | 15426.8** | 9493.2 |
| Crypt_TMIN_Lin_BS4 | Linear | NA | B-spline | 4 | No | 15427.3* | 9485.9 |
| Crypt_TMIN_Lin_BS5 | Linear | NA | B-spline | 5 | No | 15435.4 | 9486.4 |
| Crypt_TMIN_Lin_BS6 | Linear | NA | B-spline | 6 | No | 15445.1 | 9488.4 |
| Crypt_TMIN_Lin_BS7 | Linear | NA | B-spline | 7 | No | 15452.9 | 9488.5 |
| **Step 3: Predictor space model variants** |  |  |  |  |  |  |  |
| Crypt_TMIN_NS2_NS2 | N-spline | 2 | N-spline | 2 | No | 15414.8 | 9473.4 |
| Crypt_TMIN_NS3_NS2 | N-spline | 3 | N- spline | 2 | No | 15377.7*** | 9421.0 |
| Crypt_TMIN_NS4_NS2 | N-spline | 4 | N-spline | 2 | No | 15392.2* | 9420.2 |
| Crypt_TMIN_NS5_NS2 | N-spline | 5 | N-spline | 2 | No | 15410.6 | 9423.1 |
| Crypt_TMIN_PS5_NS2 | P-spline | 5 | N-spline | 2 | No | 15399.9 | 9412.5 |
| Crypt_TMIN_BS3_NS2 | B-spline | 3 | N-spline | 2 | No | 15388.5** | 9431.8 |
| Crypt_TMIN_BS4_NS2 | B-spline | 4 | N-spline | 2 | No | 15407.4 | 9435.3 |
| Crypt_TMIN_BS5_NS2 | B-spline | 5 | N-spline | 2 | No | 15410.3 | 9422.9 |
| Crypt_TMIN_DTHR_NS2 | D-Thresh | Knot = 0 | N-spline | 2 | No | 15425.8 | 9484.5 |
| Crypt_TMIN_HTHR_NS2 | H-Thresh | Knot = 80^th^ Perc. | N-spline | 2 | No | 15691.5 | 9773.2 |
| Crypt_TMIN_LTHR_NS2 | L-Thresh | Knot = 20^th^ Perc. | N-spline | 2 | No | 15726.4 | 9808.1 |
| Crypt_TMIN_NS2_BS3 | N-spline | 2 | B- spline | 3 | No | 15430.0 | 9473.3 |
| Crypt_TMIN_NS3_BS3 | N-spline | 3 | B- spline | 3 | No | 15394.9 | 9415.1 |
| Crypt_TMIN_NS4_BS3 | N-spline | 4 | B- spline | 3 | No | 15401.2 | 9398.4 |
| Crypt_TMIN_NS5_BS3 | N-spline | 5 | B- spline | 3 | No | 15426.4 | 9400.5 |
| Crypt_TMIN_PS5_BS3 | P-spline | 5 | B-spline | 3 | No | 15406.8 | 9380.9 |
| Crypt_TMIN_BS3_BS3 | B-spline | 3 | B-spline | 3 | No | 15400.6 | 9420.9 |
| Crypt_TMIN_BS4_BS3 | B-spline | 4 | B-spline | 3 | No | 15412.6 | 9409.8 |
| Crypt_TMIN_BS5_BS3 | B-spline | 5 | B-spline | 3 | No | 15415.5 | 9389.7 |
| Crypt_TMIN_DTHR_BS3 | D-Thresh | Knot = 0 | B-spline | 3 | No | 15442.2 | 9485.5 |
| Crypt_TMIN_HTHR_BS3 | H-Thresh | Knot = 80^th^ Perc. | B-spline | 3 | No | 15699.4 | 9773.4 |
| Crypt_TMIN_LTHR_BS3 | L-Thresh | Knot = 20^th^ Perc. | B-spline | 3 | No | 15714.2 | 9788.2 |
| Crypt_TMIN_NS2_BS4 | N-spline | 2 | B-spline | 4 | No | 15438.2 | 9466.1 |
| Crypt_TMIN_NS3_BS4 | N-spline | 3 | B-spline | 4 | No | 15414.0 | 9411.2 |
| Crypt_TMIN_NS4_BS4 | N-spline | 4 | B-spline | 4 | No | 15429.2 | 9395.6 |
| Crypt_TMIN_NS5_BS4 | N-spline | 5 | B-spline | 4 | No | 15463.4 | 9399.1 |
| Crypt_TMIN_PS5_BS4 | P-spline | 5 | B-spline | 4 | No | 15435.3 | 9371.0 |
| Crypt_TMIN_BS3_BS4 | B-spline | 3 | B-spline | 4 | No | 15418.7 | 9415.9 |
| Crypt_TMIN_BS4_BS4 | B-spline | 4 | B-spline | 4 | No | 15433.5 | 9399.9 |
| Crypt_TMIN_BS5_BS4 | B-spline | 5 | B-spline | 4 | No | 15446.6 | 9382.2 |
| Crypt_TMIN_DTHR_BS4 | D-Thresh | Knot = 0 | B-spline | 4 | No | 15451.6 | 9479.5 |
| Crypt_TMIN_HTHR_BS4 | H-Thresh | Knot = 80^th^ Perc. | B-spline | 4 | No | 15699.3 | 9765.7 |
| Crypt_TMIN_LTHR_BS4 | L-Thresh | Knot = 20^th^ Perc. | B-spline | 4 | No | 15723.8 | 9790.1 |
| **Step 4: Models stratified by season** |  |  |  |  |  |  |  |
| Crypt_TMIN_NS3_NS2_X | N-spline | 3 | N-spline | 2 | Yes | 15444.6 | 9349.6* |
| Crypt_TMIN_BS3_NS2_X | B-spline | 3 | N-spline | 2 | Yes | 15424.9 | 9329.8** |
| Crypt_TMIN_NS4_NS2_X | N-spline | 4 | N- spline | 2 | Yes | 15465.8 | 9309.2*** |

DF = Degrees of freedom

BIC = Bayesian information criterion

AIC = Akaike information criterion

NA = Not applicable (i.e. DF not specified)

To demonstrate the analytical process that was used in this study, all variants of our minimum temperature and cryptosporidiosis models are detailed above. After performing exploratory analyses in Step 1 (Not depicted above), Step 2 was to hold the predictor space constant with a linear function, while third degree natural spline, B-spline and penalized spline functions were compared for the lag space, incrementally increasing the degrees of freedom (DF), up to a maximum of seven. The three best cross-basis definitions for the lag space (indicated by the BIC) were then used in Step 3, while threshold functions and third-degree spline functions were assessed for the predictor space, incrementally increasing the DF for the predictor space up to a maximum of five. In Step 4, the cross-basis formulation of the three best models (i.e. lowest BIC) identified across Steps 2 and 3 were then used in three final models that stratified by season. The three models with the lowest BIC, and the three with the lowest AIC were selected as the final set of models (highlighted in gray above), which were subsequently compared on their predictive performance using a 21-fold cross-validation process.

**Appendix Table A3. Characteristics of the top six performing models for each outcome-predictor pair.**

| **Model** | **Predictor function** | **Predictor DF/Knot** | **Lag function** | **Lag DF** | **Season**  **Interaction** | **BIC** | **AIC** | **RMSE** |
| --- | --- | --- | --- | --- | --- | --- | --- | --- |
| **Cryptosporidiosis** |  |  |  |  |  |  |  |  |
| **Maximum Temperature** |  |  |  |  |  |  |  |  |
| Crypt-TMAX-Mod1 | B-spline | 3 | N-spline | 2 | No | 15385.6 | 9428.9 | 0.5044 |
| Crypt-TMAX-Mod2 | N-spline | 4 | N-spline | 2 | No | 15389.0 | 9416.9 | 0.5062 |
| Crypt-TMAX-Mod3 | N-spline | 3 | N-spline | 2 | No | 15376.0 | 9419.3 | 0.5174 |
| Crypt-TMAX-Mod4 | N-spline | 5 | B-spline | 5 | No | 15451.7 | 9348.9 | 0.5195 |
| Crypt-TMAX-Mod5 | N-spline | 4 | N-spline | 2 | Yes | 15480.1 | 9323.5 | 0.5752 |
| Crypt-TMAX-Mod6 | N-spline | 3 | N-spline | 2 | Yes | 15451.7 | 9356.6 | 0.5816 |
| **Minimum Temperature** |  |  |  |  |  |  |  |  |
| Crypt-TMIN-Mod1 | N-spline | 3 | N-spline | 2 | No | 15377.7 | 9421.0 | 0.5015 |
| Crypt-TMIN-Mod2 | B-spline | 3 | N-spline | 2 | No | 15388.5 | 9431.8 | 0.5028 |
| Crypt-TMIN-Mod3 | N-spline | 4 | N-spline | 2 | Yes | 15465.8 | 9309.2 | 0.5046 |
| Crypt-TMIN-Mod4 | N-spline | 4 | N-spline | 2 | No | 15392.2 | 9420.2 | 0.5137 |
| Crypt-TMIN-Mod5 | N-spline | 3 | N-spline | 2 | Yes | 15424.9 | 9329.8 | 0.5189 |
| Crypt-TMIN-Mod6 | B-spline | 3 | N-spline | 2 | Yes | 15444.6 | 9349.6 | 0.5393 |
| **PRECIPipitation** |  |  |  |  |  |  |  |  |
| Crypt-PRECIP-Mod1 | N-spline | 2 | N-spline | 3 | No | 15411.8 | 9455.1 | 0.5000 |
| Crypt-PRECIP-Mod2 | N-spline | 2 | N-spline | 2 | No | 15406.2 | 9464.8 | 0.5007 |
| Crypt-PRECIP-Mod3 | N-spline | 2 | B-spline | 3 | No | 15412.1 | 9455.4 | 0.5021 |
| Crypt-PRECIP-Mod4 | N-spline | 2 | N-spline | 2 | Yes | 15436.6 | 9403.0 | 0.5880 |
| Crypt-PRECIP-Mod5 | N-spline | 2 | B-spline | 3 | Yes | 15494.5 | 9399.4 | 1.5665 |
| Crypt-PRECIP-Mod6 | N-spline | 2 | N-spline | 3 | Yes | 15495.6 | 9400.6 | 1.6478 |
| **Giardia** |  |  |  |  |  |  |  |  |
| **Maximum Temperature** |  |  |  |  |  |  |  |  |
| Giard-TMAX-Mod1 | Linear | 1 | N-spline | 2 | No | 27636.2 | 21710.3 | 2.0251 |
| Giard-TMAX-Mod2 | P-spline | 5 | N-spline | 2 | Yes | 27671.5 | 21684.1 | 2.0763 |
| Giard-TMAX-Mod3 | Linear | 1 | B-spline | 3 | Yes | 27668.2 | 21665.4 | 2.0961 |
| Giard-TMAX-Mod4 | N-spline | 2 | N-spline | 2 | No | 27645.3 | 21704.0 | 2.1641 |
| Giard-TMAX-Mod5 | Linear | 1 | N-spline | 2 | Yes | 27639.4 | 21667.3 | 2.7501 |
| Giard-TMAX-Mod6 | N-spline | 2 | N-spline | 2 | Yes | 27684.7 | 21651.1 | 7380.0 |
| **Minimum Temperature** |  |  |  |  |  |  |  |  |
| Giard-TMIN-Mod1 | Linear | 1 | B-spline | 3 | No | 27620.2 | 21686.5 | 2.0129 |
| Giard-TMIN-Mod2 | Linear | 1 | N-spline | 2 | No | 27615.4 | 21689.4 | 2.0138 |
| Giard-TMIN-Mod3 | D-threshold | Knot=0 | N-spline | 2 | No | 27617.8 | 21676.4 | 2.0157 |
| Giard-TMIN-Mod4 | D-threshold | Knot=0 | B-spline | 3 | No | 27627.6 | 21670.9 | 2.0172 |
| Giard-TMIN-Mod5 | D-threshold | Knot=0 | N-spline | 3 | No | 27628.0 | 21671.3 | 2.0198 |
| Giard-TMIN-Mod6 | D-threshold | Knot=0 | N-spline | 2 | Yes | 27701.9 | 21668.3 | 2.0514 |
| **Precipitation** |  |  |  |  |  |  |  |  |
| Giard-PRECIP-Mod1 | Linear | 1 | N-spline | 2 | No | 27633.5 | 21707.5 | 2.0167 |
| Giard-PRECIP-Mod2 | N-spline | 2 | N-spline | 2 | No | 27639.5 | 21698.2 | 2.0186 |
| Giard-PRECIP-Mod3 | Linear | 1 | N-spline | 3 | No | 27643.4 | 21709.7 | 2.0196 |
| Giard-PRECIP-Mod4 | N-spline | 5 | B-spline | 3 | No | 27718.2 | 21692.3 | 2.0256 |
| Giard-PRECIP-Mod5 | N-spline | 5 | N-spline | 3 | No | 27718.9 | 21693.0 | 2.0285 |
| Giard-PRECIP-Mod6 | N-spline | 2 | N-spline | 2 | Yes | 27716.9 | 21683.4 | 7.7228 |

For each of the six outcome-predictor pairs that were assessed in this study, the functions that were selected for defining the predictor space and the lag space within the cross-basis structure(s) included in each model are detailed for the three models that had the lowest AIC, and the three models that had the lowest BIC. The set of six models selected via AIC/BIC for each outcome-predictor pair was subsequently compared using a 21-fold cross-validation process. The mean RMSE value summarizing model performance across all tuning and validation iterations was then used as an indicator of the overall predictive skill of the model. The model with the lowest mean RMSE was selected as the final model, which is denoted “Mod1” and highlighted in gray for each of the six outcome-predictor pairs detailed above.
